# Guilt by association: Plant-based foods can be incorporated into both healthy and unhealthy plant-based diet indices associated with coronary heart disease

**DOI:** 10.1101/2024.06.29.24309713

**Authors:** Yasaman Jamshidi-Naeini, Beate Henschel, James M. Shikany, David B. Allison, Andrew W. Brown

## Abstract

**Background:** One approach to test for differential associations between plant foods with health uses a scoring approach: foods categorized into animal or ‘healthy’ plant-based or ‘unhealthy’ plant-based groups to construct a plant-based diet index (PDI), healthy PDI (hPDI), and unhealthy PDI (uPDI).

**Objective:** To evaluate robustness of associations between diet indices and incident coronary heart disease (CHD) risk when recategorizing food groups in indices.

**Methods:** Using REasons for Geographic and Racial Differences in Stroke (REGARDS) data, we replicated a published use of the scoring approach. Using Cox proportional hazards regression, we assessed ramifications of the following on associations between diet indices and CHD risk: 1) reconfiguring foods within and among food groups, using potatoes as an example, 2) leave-one-out analysis for each of 12 plant-based food groups, and 3) agnostically redefining each food group as ‘healthy’ or ‘unhealthy’.

**Results:** Over 153,286 person-years of follow-up, there were 868 cases of CHD. Replication analyses did not reach statistical significance. General patterns of magnitude of hazard ratios (HRs) in replication and reconfiguration models were PDI HRs < hPDI HRs < uPDI HRs for women, and hPDI < PDI < uPDI for men. Five models reconfiguring potatoes resulted in small, varied differences in PDI, hPDI, and uPDI associations. Leave-one-out analyses resulted in greater variation of associations between indices and CHD. In agnostic models, each plant-based food group was classified in indices as ‘healthy’ and ‘unhealthy’ with statistically significant beneficial or deleterious associations with CHD. Averaged over 4,096 models, HRs’ shifts were small when food groups were moved between ‘healthy’ and ‘unhealthy’.

**Conclusion:** Statistically significant associations between hPDI, uPDI, and PDI and incident CHD were not replicated. Small perturbations of the scoring approach had varied impacts on HRs. Agnostically constructing diet indices demonstrated the potential for guilt (or benefit) by association: any of the food groups we studied could be categorized with others in an index showing beneficial or deleterious associations.

## Introduction

The distinctions between ‘healthy’ and ‘unhealthy’ (sometimes called ‘less-healthy’) plant-based foods are of much interest for diet and health researchers, practitioners, and guideline creators. Research groups have looked at the association of various interpretations of plant-based diets with different health outcomes including overweight/obesity, weight change, cognition, mortality, type 2 diabetes (T2D), overall cardiovascular disease (CVD), coronary heart disease (CHD), breast and overall cancer, biological ageing trajectory, and risk of frailty (1–11). However, the demarcations between ‘healthy’ and ‘unhealthy’ varies depending on factors including glycemic indices and glycemic loads calculated on individual foods rather than meals or diets; median sex-specific splits of self-reported consumption within cohorts; or researcher discretion on choosing what they believe are more or less ‘healthy’. Categories of foods can also vary based on component foods, energy density, numbers and varieties of foods, and other characteristics. The general approach to developing diet indices involves investigators deciding which foods they consider fitting their construct through data-driven approaches, evaluation of prior literature, or ethnographic determinations. Thereafter, researchers will assess adherence to that construct in some way. Such an approach has been used for different food consumption patterns, including Mediterranean diets and ‘plant-based’ diets.

One particular extension for plant-based foods advanced by Satija et al. was to categorize food frequency questionnaire (FFQ) data into ‘healthy’ plant-based, ‘less healthy’ plant-based, or animal food groups. The distinctions were driven by their interpretation of “existing knowledge of associations of the foods with T2D, other outcomes (CVD, certain cancers), and intermediate conditions (obesity, hypertension, lipids, inflammation)” (2). Using data from Nurses’ Health Study (NHS), NHS2, and Health Professionals Follow-Up Study (HPFS), the authors presented associations between plant diet indices and T2D (2), cardiometabolic outcomes (3), and weight change (4). Given the adoption of the approach by the original authors and others (e.g., (1–8), the properties of this approach are worth studying to assess how analytical choices may influence conclusions. This is in the spirit of the analogy advanced by Sam Savage of “shaking the ladder” to assess its stability “to minimize the risk that it falls down with you on it” (12).

### The scoring approach

The method advanced by Satija et al. (hereafter, the scoring approach) involves using FFQ data to estimate daily servings of consumption of individual foods. The individual foods are then aggregated into food groups (e.g., individual vegetables within the ‘vegetables’ food group). The sum of servings of foods within a group is then divided by quintiles and scored from 1 to 5 depending on whether a food is ‘healthy’ or ‘unhealthy’. The scores from each food group are then summed to get an index, and, in the main analysis, the index values are themselves divided by deciles. The ultimate exposure score is therefore highly transformed, and like other index-based approaches, loses much of the identity of the dietary exposure components. The scoring approach has important limitations, some of which are shared by other index-based approaches or nutrition epidemiology in general, including that estimating daily dietary intakes from self-report such as FFQ may not accurately reflect actual intakes (13–16) and removing “implausible” (i.e., extreme) energy intake reporters may not recover unbiased diet-health associations from misreporting (17). Our replication of the scoring approach has a further limitation of extrapolating from FFQ data to grams instead of just servings given the data we used (see Methods).

For the purposes of the present investigation, we set aside these limitations and instead focus on how aggregation of foods into categories in the scoring approach may influence the strength and nature of associations between foods and health outcomes. The justification of categorization schemes depends on purpose (e.g., maximizing explained variance; detecting maximal associations; determining associations with food composition; testing associations with sociological dietary constructs). Furthermore, it is improbable that a food is wholly ‘unhealthy’ or ‘healthy’ for all circumstances, people, and outcomes (18). Thus, there is likely no single ‘correct’ definition of food categories. However, understanding the effect of reclassification of foods on the associations between diet indices and health outcomes is essential to determine the robustness and stability of such associations. Throughout the text, we use quotation marks around ‘healthy’ and ‘unhealthy’ to emphasize that these labels are used for categorization, not to indicate a determination that the foods inherently have health promoting or deleterious health effects.

### Objectives

Herein, we examine how different ways of categorizing foods in the scoring approach affect the dietary associations with cardiometabolic outcomes in four ways. First, we attempt to replicate the scoring approach from Satija et al. on associations between diet indices and CHD using a different data set. Second, given that potatoes were uniquely assigned as a single-plant category among all plant-based foods, we use potatoes as a case study to investigate the robustness of the scoring approach to recategorization of foods and a food category. Third, we determine the ramifications of each of the 12 plant food groups using a leave-one-out analysis. Fourth, we use an agnostic approach by assigning plant food groups as ‘healthy’ or ‘unhealthy’ to determine the distribution of associations across 4,096 models, and assessing the differences between the 2,048 models in which a food group was classified as ‘healthy’ and the 2,048 in which it was classified as ‘unhealthy’. We hypothesized that categorizing foods in different ways (e.g., within and/or between ‘healthy’ and ‘unhealthy’ categories as modeled in the scoring approach) would have a marked impact on the associations between diet indices and combined incident nonfatal myocardial infarction (MI) and fatal CHD (together referred to as CHD herein).

### Methods

We replicated to the best of our ability the analyses from Satija et al. (3) using the REasons for Geographic and Racial Differences in Stroke (REGARDS) data, including cohort creation (eligibility criteria), outcome ascertainment, selection of adjustment covariates, and other methodologic choices. This allowed us to focus on the effects of changes to the scoring approach within a single statistical associational framework.

### Study population

As described elsewhere, REGARDS is an ongoing population-based longitudinal study in 30,239 randomly sampled adults from all 48 continental US states to examine reasons for regional and racial differences in stroke mortality (19, 20). The REGARDS study was previously approved by all associated institutional review boards (19). Briefly, between January 2003 and October 2007, participants (Black or White race; aged ≥ 45 years) completed a 45-min computer-assisted telephone interview on socio-demographics, medical history, and health status followed by an in-home examination to collect fasting blood and urine samples, electrocardiograms (ECG), and data on blood pressure measures, anthropometric measures, and medications taken in the 2 weeks before the visit via pill bottle review. An FFQ was left for self-administration. REGARDS participants are contacted every six months by phone and interviewed about stroke symptoms, hospitalizations, and general health status (19). . By the time of the current analysis, the REGARDS study had conducted two in-home visits for data collection: one at baseline and a follow-up visit approximately 10 years later. The data collection during the second in-home visit was similar in terms of the variables on which data were collected to the first visit.

In the current analysis, we excluded individuals with history of self-reported MI or atrial fibrillation, coronary artery bypass graft, angioplasty, or evidence of MI via ECG; cancer^1^; stroke or transient ischemic attack; and missing or implausible energy intake (described in the scoring approach as <600 or >3,500 kcal/day for women and <800 or >4,200 kcal/day for men). We additionally excluded participants with missing covariate data at baseline (Supplementary Figure 1). Our final analytic sample included 13,684 individuals, 8,255 of whom had additional data from the follow-up visit.

### Adjustment covariates

To emulate Satija et al., we used the ‘updated’ covariate data from the second (i.e., most recent) REGARDS data collection for participants for whom those data were available, and baseline covariate data for the rest. Adjustment covariates include age at baseline (in years), updated multivitamin use^2^ (yes, no), family history of MI (yes, no), updated margarine intake (by quintiles^3^), updated energy intake (by quintiles), diabetes^4^ at baseline (yes, no), hypercholesterolemia at baseline^5^ (yes, no), hypertension^6^ at baseline (yes, no), updated smoking status (never, past, current (1 to <15 cigarettes per day), current (15-<25 cigarettes per day), current (25 or more cigarettes per day)), updated physical activity (none, 1-3 times per week, 4 or more times per week), updated alcohol intake^7^, updated aspirin use (yes, no), updated body mass index (BMI) category (BMI<21, 21 to <23, 23 to <25, 25 to <27, 27 to <30, 30 to <33, 33 to <35, 35 to <40, and 40 kg/m^2^ or higher), race (Black, White) and updated REGARDS region of residence (stroke belt, stroke buckle, non-belt region^8^). Race and region are specific to REGARDS and not included in the Satija et al. analysis. In the analysis stratified by gender, we also adjusted for hormone use (never, past, current) and past oral contraceptive use (yes, no) among female participants.

### Outcome ascertainment

CHD was defined as incident nonfatal MI and fatal CHD. In the REGARDS dataset, fatal CHD was defined as “death within 28 days of an adjudicated definite or probable MI, or CHD or sudden death as the adjudicated cause of death.[…] The main underlying cause of death was determined by 2 trained adjudicators who examined all available information including interviews with next of kin, death certificates, autopsy reports, medical history, and the National Death Index. […] [In REGARDS, nonfatal MI ascertainment] required a clinical presentation consistent with ischemia, a rising and falling pattern of troponin over at least 6 hours with a peak at least twice the upper limit of normal, or imaging findings consistent with ischemia (20).” Events up to December 31, 2019 were available for the current analysis.

### Dietary assessment

REGARDS dietary and alcohol data were collected from a self-administered 110-item Block 98 FFQ (NutritionQuest, Berkeley, CA) given to participants during in-home visits to complete and return by mail to the REGARDS Operations Center, where they were reviewed for completeness, scanned, and forwarded to NutritionQuest for processing and analysis (21, 22). The dataset we received had frequency and amount of consumption of each FFQ item transformed into consumed grams per day.

#### o Replicating the scoring approach food groups using REGARDS FFQ items

We assigned each REGARDS FFQ item to one of 18 food groups defined by the scoring approach (2, 3). We relied on the details in the Satija et al. publications to determine the items that composed each of the 18 food groups, resulting in three situations: 1) A REGARDS FFQ item was matched with an identical item of the food groups in the scoring approach (e.g., broccoli, dark bread, strawberries, eggs, hot dog, yogurt, coffee); 2) A REGARDS FFQ item was not identically listed in Satija et al. (2, 3), but based on the available details and 1984 NHS FFQ, the FFQ item clearly fit into one of the food groups. For instance, Raisin Bran cereal, Chinese dish, and different types of chicken from REGARDS did not have an exact match, but could be reasonably fit under the food groups whole grains, miscellaneous animal-based foods, and meat, respectively; and 3) A REGARDS FFQ item could not be matched with a specific item, nor did it clearly fit into one of the food groups, and were thus excluded (e.g., breakfast bars, gravy, non-dairy creamer, water, salsa). The scoring approach used cumulatively averaged dietary data over the follow-up durations of NHS, NHS2 and HPFS. We thus averaged values of the two REGARDS data points (i.e., first and second visits)for dietary information if dietary data from the second visit were available, unless type 2 diabetes was diagnosed before the second data capture because this may have resulted in substantial dietary change. When follow-up data were not available, baseline dietary data were carried forward.

#### o Plant-based diet indices in the scoring approach

To calculate PDI, hPDI, and uPDI of the scoring approach, Satija and colleagues (2, 3) calculated the sum of foods in a food group, created quintiles for these summed intakes of food groups, and assigned a score ranging from 1 to 5 per food group per participant. For positive scoring, participants received higher scores for higher reported intakes and vice versa. For reverse scoring, participants received lower scores for higher reported intakes. Scoring schemes for calculating PDI, hPDI, and uPDI are presented in Table 1. To obtain the PDI, hPDI, and uPDI scores, the scores of all 18 food groups were summed. Thus, all three indices range in theory from 18 to 90.

**Table 1.** Scoring schemes for creating plant-based diet indices.

|  | PDI | hPDI | uPDI |
| --- | --- | --- | --- |
| Healthy Plant Food Groups |  |  |  |
| Whole grains | Positive | Positive | Reverse |
| Fruits |  |  |  |
| Vegetables |  |  |  |
| Nuts |  |  |  |
| Legumes |  |  |  |
| Vegetable oils |  |  |  |
| Tea and coffee |  |  |  |
| Less Healthy Plant Food Groups |  |  |  |
| Fruit juices | Positive | Reverse | Positive |
| Refined grains |  |  |  |
| Potatoes |  |  |  |
| Sugar sweetened beverages |  |  |  |
| Sweets and desserts |  |  |  |
| Animal Food Groups |  |  |  |
| Animal fat | Reverse | Reverse | Reverse |
| Dairy |  |  |  |
| Egg |  |  |  |
| Fish or seafood |  |  |  |
| Meat |  |  |  |
| Miscellaneous animal-based foods |  |  |  |

### Statistical analysis

We replicated the analytical procedures as previously reported as closely as possible (2, 3). Briefly, we used Cox proportional hazards regression to estimate hazard ratios (HR) and 95% confidence intervals (CIs) evaluating the associations of continuous index scores and deciles of each index separately with incident CHD. Person-time was calculated from age at questionnaire return date until CHD diagnosis, death, or end of follow-up (December 31, 2019). We used age (in years) as the time scale, but we were not able to stratify by calendar time as done by Satija and colleagues because of shorter follow-up. Indices were averaged over the two dietary data captures except for the cases with the diagnosis of T2D. Models were adjusted for the aforementioned covariates.

For completeness in replication, we conducted p-for-trend analyses by assigning median values to each decile bin (e.g., bin 1 from below decile 1; bin 10 from above decile 9), and this variable was used in the analysis. These analyses and results are presented in the supplementary materials.

Satija et al.’s analysis was in practice stratified by sex because they used sex-specific cohorts (women: NHS and NHS2; men: HPFS). Thus, we used gender in REGARDS data as a stratification variable. As in Satija et al., we combined the stratified results using a fixed-effects meta-analysis. Variable recoding was done using SAS software version 9.4 (SAS Institute Inc., Cary, North Carolina) and all analyses were performed in R version 4.2.3 and RStudio version 2023.03.0 (R Core Team (2023). R: A language and environment for statistical computing. R Foundation for Statistical Computing, Vienna, Austria. https://www.R-project.org/.). Statistical significance was set at a 2-tailed p-value of <0.05.

### Recategorization schemes for potatoes

In the scoring approach ‘French fries’, ‘baked or mashed potatoes’, and ‘potato or corn chips’ are grouped as ‘potatoes’ as an ‘unhealthy’ food group. Our replicated ‘potatoes’ group included white potato^9^, French fries^10^, salty snacks^11^, and low-fat salty snacks^12^. Much like the scoring approach, we included FFQ items that were not exclusively potatoes if the item was predominantly potatoes, despite creating a bit of a misnomer in the ‘potatoes’ group. To examine the influence of food categorization schemes on associations between diet indices and CHD, we used five alternative categorization schemes by moving all or some of these items within or between ‘healthy’ and ‘unhealthy’ food groups. We then re-calculated the PDI, hPDI, and uPDI for each recategorization and estimated HRs and 95% CIs for each model. Thus, we have results from six models: the scoring approach replication and five alternative models. Table 2 shows the five alternative approaches that we used to categorize potato food items. When the number of food groups was either increased (model 4) or decreased (models 1 and 3) we rescaled the diet indices to match the original scale that ranged from 18 to 90 points. Note that the PDI scores from model 2 are identical to the scoring approach because in calculating PDI all plant food groups are scored positively.

**Table 2.** Recategorization Approaches for ‘Potatoes’ food group^1^.

| Recategorization approach | Description |
| --- | --- |
| Model 1: "exclude potato group" | Exclude all items in “potatoes” group from indices |
| Model 2: “potato group in healthy” | Move “potatoes” food group as a whole and unchanged group into “Healthy” category |
| Model 3: "potatoes in vegetable group" | Move all items in “potatoes” group into “vegetables” group |
| Model 4: "healthy and unhealthy potatoes" | Split up “potatoes” group:<br>- Move white potato as a single item group into “Healthy” category,<br>- Keep French fries, salty snacks, and low-fat salty snacks in “potatoes” under “less-healthy” category |
| Model 5: "vegetables and unhealthy potatoes" | Split up “potatoes” group:<br>- Move white potato into “Vegetables” group,<br>- Keep French fries, salty snacks, and low-fat salty snacks in “Potatoes” under “less-healthy” category |

<sup>1</sup> Our replication of the ‘potatoes’ food group using the REGARDS data consists of white potato, French fries, salty snacks, and low-fat salty snacks.

All food groups categorized under plant foods (either ‘healthy’ or ‘unhealthy’) are scored positively in the PDI. Thus, it is expected that associations of PDI with CHD should remain largely unchanged in the recategorization approach. “Potatoes” as a food group is categorized under the ‘unhealthy’ category in the scoring approach and scored in reverse in hPDI calculation (i.e., higher consumption of potato food items leads to lower hPDI scores). If one assumes that items in the “potatoes” food group have a meaningful contribution to any detectable association with the risk of CHD, and if all or some of the items in the “potatoes” group are miscategorized as ‘unhealthy’ when in fact they should be ‘healthy’, moving them to ‘healthy’ should result in stronger, inverse associations (lower HRs) between hPDI and CHD risk (that is: as hPDI scores go up, CHD risk goes down). Similarly, for uPDI, if potatoes are incorrectly categorized as ‘unhealthy’ when in fact they should be categorized as ‘healthy’, moving all or some of them to ‘healthy’ should make the associations between uPDI and CHD risk stronger. As such, hPDI is expected to have the strongest beneficial association followed by PDI, while uPDI is expected to have the strongest harmful association with CHD (hPDI<PDI<uPDI).

### Leave-One-Out Analyses

PDI, hPDI, and uPDI scores were calculated as done in the scoring approach but leaving one of the 12 plant-based food groups out of the calculation at a time. This analysis aims to determine the influence of perturbing an entire food group on the model results, which reveals the importance of each plant-based food group as well as creating a benchmark against which to compare the magnitude of the recategorization results. When potatoes were left out, this approach is equivalent to Model 1 from the recategorization approach above.

### Agnostic permutation analyses

Indices were calculated using all possible combinations of each of the 12 plant-based food groups scored either positively or reversely (2^12=4096 combinations). Because uPDI and hPDI are coded oppositely, and all combinations were tested, uPDI results mirror the results of hPDI. Therefore, we only present the combinations as hPDI. We examined the distribution of HRs from positive and reverse scoring using paired t-tests of the log HRs (separately for each plant-based food group).

### Results and Interpretations

During 153,286 person-years of follow-up, there were 868 cases of incident CHD (388 women with 92,334 person-years and 480 men with 60,952 person-years).

### Replicating the Scoring Approach of Satija et al

We obtained comparable median values within decile bins of PDI, hPDI, and uPDI in the replicated models compared with Satija et al.’s median values (3) (see Supplementary Table 1). For instance, men’s hPDI median value was 42.0 in decile bin 1 and 65.0 in bin 10 in our replicated calculation, which compares with 43.0 in bin 1 and 66.0 in bin 10 in (3). We did not observe statistically significant associations of PDI and uPDI scores and the risk of CHD for women, men, or both combined (see Supplementary Tables 2 and 4). Most associations for hPDI scores and risk of CHD were also not statistically significant; only the following HRs were significant at p<0.05: decile bin 7 for women and combined, bin 9 for men, bin 5 for combined, p for trend in combined (see Supplementary Table 3).

### Reconfiguring potatoes within the scoring approach

Supplementary Table 1 displays the median values for each decile bin of PDI, hPDI, and uPDI scores by gender obtained in the reconfiguration approaches from Table 2. Because scores are constrained by quantiles (i.e., if one individual’s score goes up another must come down), differences in the scores were small (<2 in any decile category of index and configuration). Comparing the mean diet scores among the five reconfigurations (see Supplementary Figure 2), we similarly see only minimal changes (<0.2 within or between patterns and configurations).

In total, the use of deciles resulted in 9 comparisons (decile bin 2 to bin 1; bin 3 to bin 1, etc…), plus the continuous analysis per 10-unit change in diet index score, and a ‘p-for-trend’ analysis that uses means of decile bins in a single regression. All decile comparisons are reported for transparency (Supplementary Tables 2-4), but for ease of interpretation, and consistent with the concept of dose-response and monotonic effect, only the extreme decile comparison will be discussed of the decile comparisons, in addition to the continuous analyses. Point estimates of HRs were qualitatively evaluated as moving towards or away from the null (null association shown as HR=1.00) compared with the scoring approach. A lower HR for hPDI and a higher HR for uPDI, as compared with the scoring approach, suggest that the reconfigured models show stronger associations. Stronger associations are consistent with recategorization of potato food items more effectively capturing the concept of ‘healthy’ and ‘unhealthy’ food groups.

Figure 1 shows the associations between PDI, hPDI, or uPDI (HRs, continuous per 10-unit change in diet indices in upper panel; highest versus lowest decile in lower panel) with CHD risk in the five recategorization schemes for potatoes by the p-value of the HRs. Points moving in the horizontal direction (compared with the replicated scoring approach) indicate a change in the strength of the association, while points moving in the vertical direction indicate a change in the significance level.

**Figure 1:**
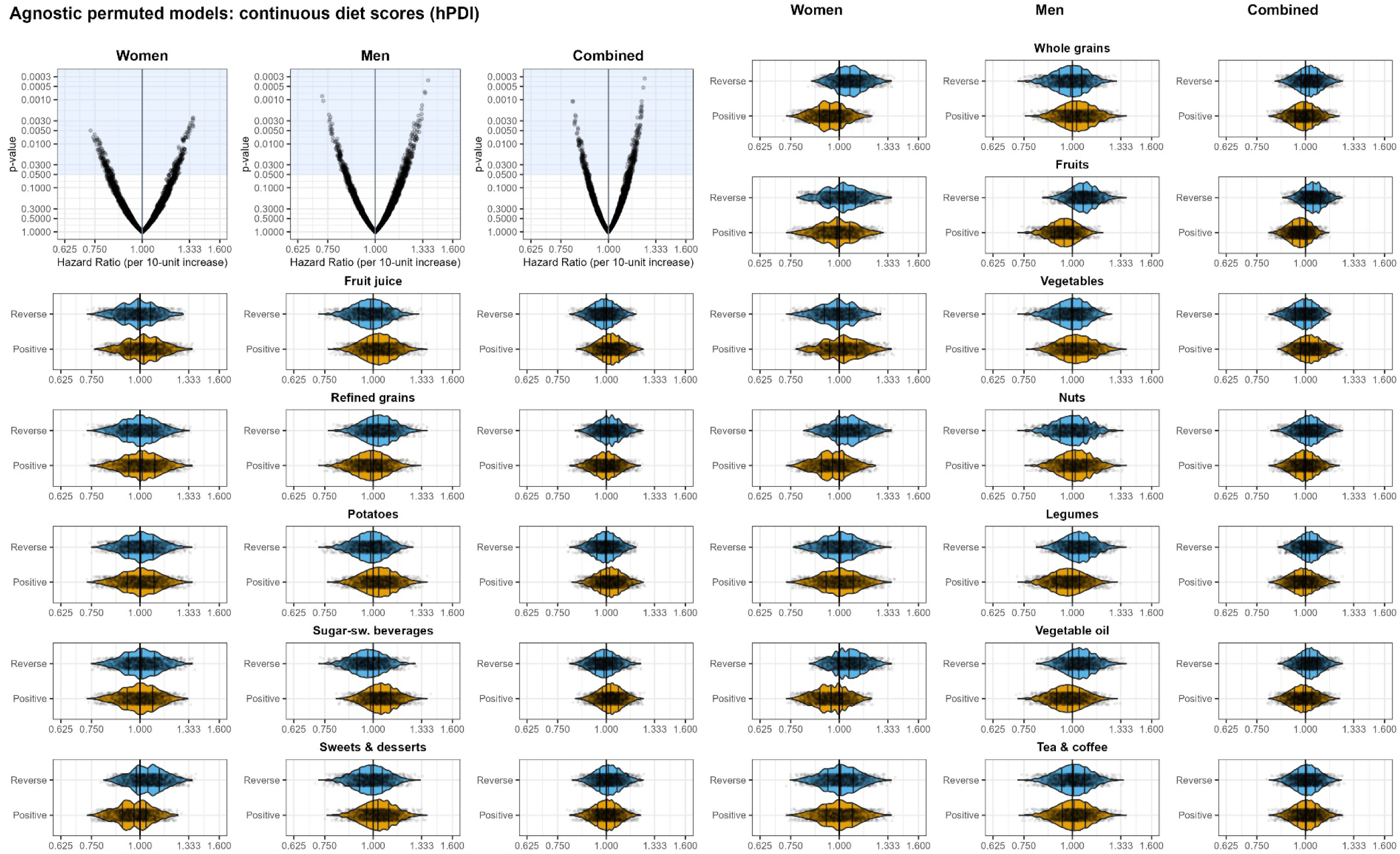
Associations of PDI, hPDI, and uPDI indices and CHD for five different recategorization schemes for potatoes (see Table 2) and our replication of the scoring approach. Top panel: per 10 units using continuous diet scores. Bottom panel: diet scores as deciles, shown are HR for highest decile compared with lowest decile. For specific numerical values, refer to Supplementary Table 2 (for PDI), Supplementary Table 3 (for hPDI), and Supplementary Table 4 (for uPDI). Black – (Scoring approach (as in Satija): Replication of the Scoring Approach; Blue – “exclude potato group” (Model 1): Exclude all items in “Potatoes” group from indices; Red – “potato group in healthy” (Model 2): Move “Potatoes” group as a whole intact group into “Healthy” super category; Green – “potatoes in vegetable group” (Model 3): Move all items in “Potatoes” group into “Vegetables” group; Purple – “healthy and unhealthy potatoes” (Model 4): Split “Potatoes” group by moving white potatoes as a single item group into “Healthy” category and keeping French fries, salty snacks, and low-fat salty snacks in “Potatoes” under “less-healthy” category; Orange – “vegetables and unhealthy potatoes” (Model 5): Split “Potatoes” group by moving white potatoes into “Vegetables” group and keeping French fries, salty snacks, and low-fat salty snacks in “Potatoes” under “less-healthy” category.

Overall, the recategorization approaches resulted in minimal changes in continuous HRs (per increments of 10-units in diet scores). For instance, HRs for all five models with PDI in women were 0.82 with almost identical 95% CIs. In the analysis for men, HRs in the recategorization moved slightly towards the null (HR=1) with increasing p-values for hPDI and uPDI scores, but not for PDI scores. Thus, we are not able to make strong quantitative distinctions because there were only slight differences in precision among models, with even smaller differences in HRs. Comparing patterns overall, higher PDI and hPDI scores were consistent with lower CHD risk, and a higher uPDI score was consistent with higher CHD risk in women, which is consistent with the results reported by Satija et al., albeit not statistically significant. However, the HRs of PDI for women are lower than those of hPDI, in contrast to Satija et al.

Recategorization resulted in greater HR dispersion when using the diet scores as deciles (Figure 1, bottom panels), but the results were not fully consistent between men and women. For example, HRs for uPDI scores moved away from the null for men, while they moved towards the null for women in the recategorization approach compared with the scoring approach, with model 3 (all potato items into vegetable food group) reaching statistical significance. Comparing patterns overall, higher PDI and hPDI scores again were consistent with lower CHD risk, and a higher uPDI score was consistent with higher CHD risk in women, which is again consistent with the results reported by Satija et al., albeit typically not statistically significant. In contrast again, the HRs of PDI for women are lower than those of hPDI.

### Leave-one-out analyses

We conducted a leave-one-out analysis to determine the impact of removing other entire food groups on associations of diet indices with CHD.

Supplementary Figure 3 shows the baseline mean values of diet indices in leave-one-out models for women and men, and Supplementary Table 5 shows median values of the decile bins of PDI, hPDI and uPDI by gender and model variations in the leave-one-out approach. Similar to the reconfiguration approach, differences in the scores were small across leave-one-out patterns because of constraints of the quantile approach.

Figure 2 shows the multivariable adjusted associations of 10-unit changes (top panel) and highest vs lowest decile (bottom panel) in PDI, hPDI, and uPDI scores with incident CHD, leaving each of the 12 plant-based food groups out one at a time. For specific numerical values, see Supplementary Tables 6 (for PDI), 7 (for hPDI), and 8 (for uPDI). The general pattern of the magnitude of HRs (PDI HRs < hPDI HRs < uPDI HRs for women, hPDI < PDI < uPDI for men) was similar to the recategorizing approach and our replication of the scoring approach. Similar to the recategorization models (as shown in Figure 1), changes in magnitude of point estimates compared with the original scoring approach (depicted in black) are relatively small for continuous diet scores and much more spread out for the diet scores by deciles (Figure 2). The directional change in the point estimates (i.e., HRs) when leaving one out compared with our replication of the scoring approach varied. For instance, removing the food groups of vegetables or fruit juices resulted in the strongest beneficial associations between PDI continuous diet scores and CHD among women, with p-values below 0.05 (Figure 2, top panel). Conversely, all leave-one-out for the uPDI diet scores by deciles attenuated associations for women while for men associations were strengthened compared with the scoring approach (bottom panel).

**Figure 2:**
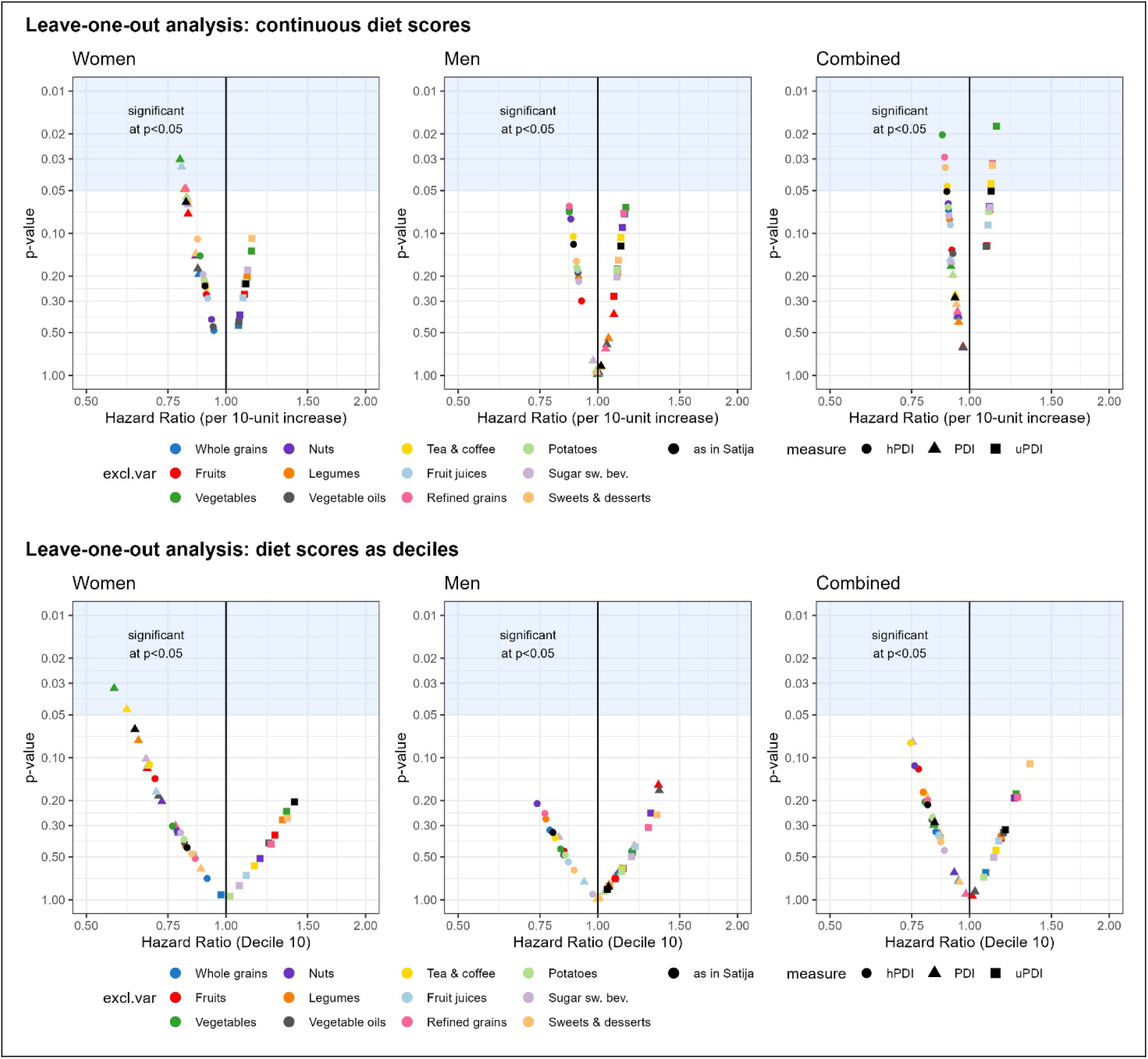
Association of PDI, hPDI, and uPDI indices and CHD for leave-one-out models. Top panel: per 10 units increment using continuous diet scores. Bottom panel: diet scores by decile, shown are HR for highest decile compared with lowest decile.

### Agnostic permutation of foods

The appropriateness of categorizing certain foods as ‘healthy’ or ‘unhealthy’ raises questions, particularly considering the shifts in associations seen with the leave-one-out approach. We therefore agonistically redefined each food group as ‘healthy’ (positively coded) or ‘unhealthy’ (reversely coded) for all food groups, resulting in 4,096 hPDI models (and 4,096 uPDI models that are exact complements; thus, only hPDIs are shown here for simplicity).

If the scoring approach is appropriately specified, then changing a food group that is actually healthy and labeled as ‘healthy’ to label it as ‘unhealthy’ (or the converse for unhealthy food groups), would be expected to weaken the associations (that is, move the hPDI toward the null HR of 1). We did not observe a consistent pattern of changes in the expected direction when ‘healthy’ or ‘unhealthy’ foods were recategorized. Instead, we observed a mix of changes in both directions and, in many cases, little difference at all. Tables 3 and 4 show comparison statistics for the distribution of agnostically defined ‘healthy’ and ‘unhealthy’ food groups, respectively. Food groups for which the positive and reverse coding resulted in a shift in HR in the expected direction for the association between hPDI score and CHD are highlighted in gray. For women, HRs were on average below 1 when whole grains, fruits, nuts, and vegetables were positively scored, indicating that higher consumption of foods in those food groups were associated with lower risk of CHD. When those same four food groups were reversely scored, the HRs were on average larger than 1, which means that lower consumption of foods in those food groups were associated with higher risk of CHD. The same trend was observed in the male subsample for fruits, legumes, and vegetable oil. Sweets and desserts for women is an example where the shifts of HRs were not in the expected direction. Scoring sweets and desserts positively (higher consumption, higher hPDI score) resulted in HRs that were smaller than 1 (i.e., higher consumption associated with lower risk of CHD). In the reverse scoring (higher consumption, lower hPDI score), HRs were on average larger than one (i.e., lower consumption associated with higher risk of CHD).

**Table 3:** Summary statistics on agnostically permuted models (hPDI score) – healthy food groups.

|  | Whole grains |  | Fruits |  | Vegetables |  | Nuts |  | Legumes |  | Vegetable Oil |  | Tea & coffee |  |
| --- | --- | --- | --- | --- | --- | --- | --- | --- | --- | --- | --- | --- | --- | --- |
|  | Positive | Reverse | Positive | Reverse | Positive | Reverse | Positive | Reverse | Positive | Reverse | Positive | Reverse | Positive | Reverse |
| **Females** |  |  |  |  |  |  |  |  |  |  |  |  |  |  |
| HR: Mean (empirical 95% spread) | 0.95<br>(0.79, 1.13) | 1.07<br>(0.90, 1.27) | 0.99<br>(0.80, 1.21) | 1.02<br>(0.83, 1.26) | 1.03<br>(0.82, 1.26) | 0.98<br>(0.80, 1.19) | 0.96<br>(0.79, 1.16) | 1.05<br>(0.87, 1.27) | 1.00<br>(0.81, 1.24) | 1.01<br>(0.82, 1.22) | 0.95<br>(0.79, 1.14) | 1.06<br>(0.89, 1.27) | 1.00<br>(0.81, 1.23) | 1.01<br>(0.82, 1.23) |
| Median (Min, Max) | 0.94<br>(0.73, 1.21) | 1.07<br>(0.85, 1.36) | 0.99<br>(0.73, 1.30) | 1.02<br>(0.78, 1.36) | 1.03<br>(0.77, 1.36) | 0.99<br>(0.73, 1.28) | 0.96<br>(0.73, 1.24) | 1.05<br>(0.81, 1.36) | 1.00<br>(0.73, 1.36) | 1.01<br>(0.76, 1.36) | 0.95<br>(0.73, 1.21) | 1.06<br>(0.84, 1.36) | 1.00<br>(0.73, 1.36) | 1.01<br>(0.75, 1.36) |
| Difference<sup>a</sup> in means | -0.12 |  | -0.04 |  | 0.04 |  | -0.09 |  | -0.00 |  | -0.11 |  | -0.01 |  |
| Ratio<sup>b</sup> of means | 0.89 |  | 0.96 |  | 1.04 |  | 0.91 |  | 1.00 |  | 0.90 |  | 0.99 |  |
| t-test<sup>c</sup> | p<0.001 |  | p<0.001 |  | p<0.001 |  | p<0.001 |  | p<0.001 |  | p<0.001 |  | p<0.001 |  |
| **Males** |  |  |  |  |  |  |  |  |  |  |  |  |  |  |
| HR: Mean (empirical 95% spread) | 1.03<br>(0.85, 1.24) | 0.99<br>(0.81, 1.17) | 0.95<br>(0.81, 1.10) | 1.07<br>(0.91, 1.24) | 1.02<br>(0.85, 1.24) | 0.99<br>(0.81, 1.17) | 1.02<br>(0.84, 1.23) | 1.00<br>(0.81, 1.20) | 0.97<br>(0.81, 1.17) | 1.04<br>(0.88, 1.24) | 0.98<br>(0.81, 1.17) | 1.04<br>(0.86, 1.23) | 1.01<br>(0.83, 1.22) | 1.00<br>(0.83, 1.21) |
| Median (Min, Max) | 1.03<br>(0.76, 1.38) | 0.99<br>(0.72, 1.30) | 0.95<br>(0.72, 1.20) | 1.07<br>(0.84, 1.38) | 1.02<br>(0.76, 1.38) | 1.00<br>(0.72, 1.26) | 1.02<br>(0.75, 1.38) | 1.00<br>(0.72, 1.35) | 0.97<br>(0.72, 1.31) | 1.04<br>(0.81, 1.38) | 0.98<br>(0.72, 1.32) | 1.04<br>(0.77, 1.38) | 1.01<br>(0.73, 1.38) | 1.01<br>(0.72, 1.35) |
| Difference<sup>a</sup> in means | 0.04 |  | -0.12 |  | 0.03 |  | 0.02 |  | -0.07 |  | -0.06 |  | 0.00 |  |
| Ratio<sup>b</sup> of means | 1.04 |  | 0.89 |  | 1.03 |  | 1.02 |  | 0.94 |  | 0.95 |  | 1.00 |  |
| t-test<sup>3</sup> | p<0.001 |  | p<0.001 |  | p<0.001 |  | p<0.001 |  | p<0.001 |  | p<0.001 |  | p<0.001 |  |

Note: Gray background highlights food groups with expected direction of associations where ‘healthy’ food groups had HR in the positive scoring (higher consumption, higher hPDI score) below 1 and HRs in the reverse scoring (higher consumption, lower hPDI score) of larger than 1; <sup>a</sup>difference = $\exp(\text{mean of } \log(\text{HR}_{\text{positive}})) - \exp(\text{mean of } \log(\text{HR}_{\text{reverse}}))$ ; <sup>b</sup>ratio = $\exp(\text{mean of } \log(\text{HR}_{\text{positive}})) / \exp(\text{mean of } \log(\text{HR}_{\text{reverse}}))$ ; <sup>c</sup>Paired t –test comparing $\log(\text{HR})$ .

**Table 4:** Summary statistics on agnostically permuted approach (hPDI score) – unhealthy food groups.

|  | Fruit juices |  | Refined grains |  | Potatoes |  | Sugar sweetened beverages |  | Sweets and desserts |  |
| --- | --- | --- | --- | --- | --- | --- | --- | --- | --- | --- |
|  | Positive | Reverse | Positive | Reverse | Positive | Reverse | Positive | Reverse | Positive | Reverse |
| **Females** |  |  |  |  |  |  |  |  |  |  |
| HR: Mean (empirical 95% spread) | 1.02 (0.83, 1.25) | 0.99 (0.80, 1.21) | 1.01 (0.82, 1.25) | 1.00 (0.81, 1.22) | 1.00 (0.81, 1.24) | 1.01 (0.82, 1.22) | 1.00 (0.81, 1.22) | 1.01 (0.82, 1.24) | 0.96 (0.79, 1.16) | 1.05 (0.87, 1.27) |
| Median (Min, Max) | 1.02 (0.76, 1.36) | 0.99 (0.73, 1.29) | 1.01 (0.74, 1.36) | 1.00 (0.73, 1.34) | 1.01 (0.73, 1.36) | 1.01 (0.75, 1.36) | 1.00 (0.73, 1.33) | 1.01 (0.75, 1.36) | 0.97 (0.73, 1.25) | 1.05 (0.81, 1.36) |
| Difference<sup>a</sup> in means | 0.04 |  | 0.01 |  | -0.00 |  | -0.01 |  | -0.08 |  |
| Ratio<sup>b</sup> of means | 1.04 |  | 1.01 |  | 1.00 |  | 0.99 |  | 0.92 |  |
| t test<sup>c</sup> | p<0.001 |  | p<0.001 |  | p<0.001 |  | p<0.001 |  | p<0.001 |  |
| **Males** |  |  |  |  |  |  |  |  |  |  |
| HR: Mean (empirical 95% spread) | 1.03 (0.85, 1.23) | 0.98 (0.81, 1.18) | 0.98 (0.81, 1.18) | 1.04 (0.86, 1.23) | 1.03 (0.85, 1.24) | 0.98 (0.81, 1.16) | 1.05 (0.88, 1.24) | 0.97 (0.81, 1.16) | 1.03 (0.85, 1.23) | 0.99 (0.81, 1.19) |
| Median (Min, Max) | 1.03, 0.77, 1.38 | 0.98, 0.72, 1.31 | 0.98, 0.72, 1.33 | 1.03, 0.77, 1.38 | 1.03, 0.76, 1.38 | 0.99, 0.72, 1.26 | 1.05, 0.80, 1.38 | 0.97, 0.72, 1.28 | 1.03, 0.76, 1.38 | 0.99, 0.72, 1.32 |
| Difference<sup>a</sup> in means | 0.05 |  | -0.05 |  | 0.05 |  | 0.08 |  | 0.04 |  |
| Ratio<sup>b</sup> of means | 1.05 |  | 0.95 |  | 1.05 |  | 1.08 |  | 1.04 |  |
| t test<sup>c</sup> | p<0.001 |  | p<0.001 |  | p<0.001 |  | p<0.001 |  | p<0.001 |  |

Note: gray background highlights food groups with expected direction of associations where ‘unhealthy’ food groups had HR in the positive scoring (higher consumption, higher hPDI score) larger than 1 and HR in the reverse scoring (higher consumption, lower hPDI score) of below 1; <sup>a</sup>difference = exp(mean of log(HR<sub>positive</sub>)) – exp(mean of log(HR<sub>reverse</sub>)); <sup>b</sup>ratio = exp(mean of log(HR<sub>positive</sub>)) / exp(mean of log(HR<sub>reverse</sub>)). <sup>c</sup>Paired t test comparing log(HR).

Violin plots in Figure 3 display the distribution of HRs for the association of continuous hPDI scores with CHD risk, with the distribution of HRs for the five ‘unhealthy’ food groups from the scoring approach on the left side and the seven ‘healthy’ food groups on the right side. hPDI scores by deciles (Supplementary Figure 4) were qualitatively similar to the results from continuous hPDI scores. Empirical cumulative density functions of the associations from Figure 3 are shown in Supplementary Figures 5 and 6 to visualize differences in distributions between positive and reverse coding of each food group.

**Figure 3:**
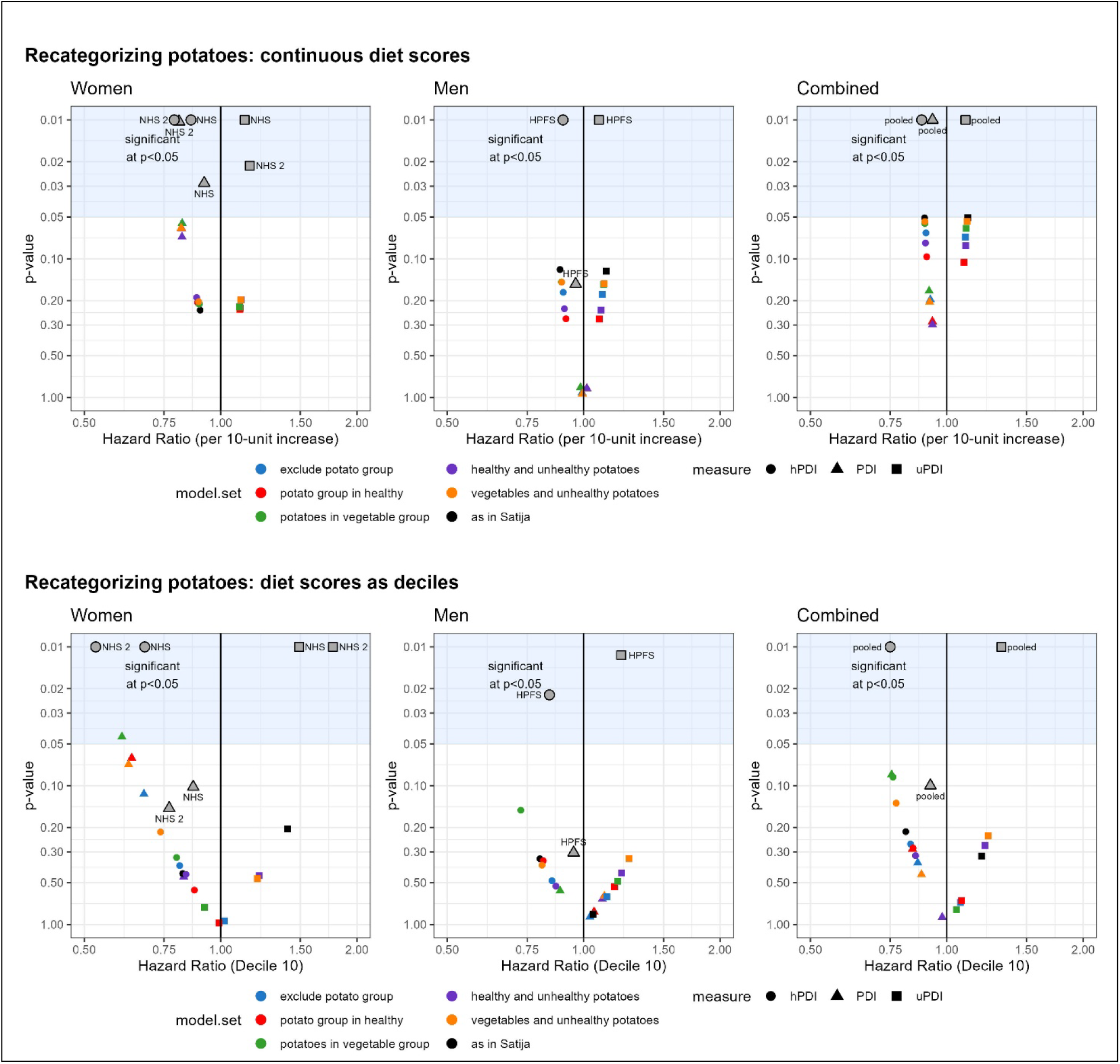
Agnostically permuted models of 12 plant-based food groups. Each of the 12 plant-based food groups are either positively (i.e., more consumption is better; depicted in orange) or reversely (less consumption is better, depicted in blue) scored, using all combinations for a total of 4,096 models. The upper left volcano plots show all 4,096 models. Each panel within sex represents the same set of models but stratified by whether that particular food group is positively or reversely coded. The 5 food groups on the left side were originally coded as ‘unhealthy’ in the scoring approach and the 7 food groups on the right side were coded as ‘healthy’. Comparisons of distributions are shown in Tables 3 and 4.

## Discussion

In this methodological exploration of a plant-based dietary scoring approach, 1) we attempted to replicate associations between plant-based diet indices and risk of CHD; 2) we adopted a pragmatic approach in rearranging the foods within and among categories to test the robustness of associations between diet indices and CHD risk, using potatoes as a particular case study; 3) we tested the ramifications of leaving each of the 12 plant-based food groups out of the calculation one at a time; and 4) we calculated hPDI scores using all possible combinations of each of the 12 plant-based food groups scored either positively or reversely (i.e., agnostically assigning plant-based food groups as ‘healthy’ or ‘unhealthy’), and compared the distributions.

Our replications of the scoring approach were not statistically significant, but were generally similar in magnitude to the results of Satija et al.; that is, the plant diet index associations were of small magnitude, but generally in the same direction, with the exception of PDI resulting in more extreme HR point estimates than the hPDI for women. We acknowledge, however, that the REGARDS dataset is smaller than the datasets used by Satija et al. (3), and our study is not an exact replication of their study. Nonetheless, the focus of our study was to explore robustness and performance of the scoring approach, rather than to draw conclusions regarding actual associations of healthfulness of foods and food groups given the other limitations of the approach. PDI scores did not differ much across the models that reconfigured potatoes as a case study (i.e., models 1-5). This is not surprising based on the way the indices were constructed using a quintile-decile approach; the approach focuses on relative rankings of individuals by quintiles of consumption rather than the specific foods consumed. Thus, moving foods within and among categories (as done in models 1-5) may not appreciably alter the overall diet scores. Evaluating the healthfulness of individual food groups using results from indices, as opposed to providing inferences in terms of the intact index, can contribute to confusion regarding the relationship between food and disease. For instance, an index from the scoring approach very well may support an association with CHD; however, support for conclusions regarding any one food group within the index may be inappropriate. As an example, ‘sweets and desserts’ showed more beneficial associations with CHD in women, consistent with the discussion about “Nutrition Science’s Most Preposterous Result” (23); yet, the differences were on par with the deleterious associations with CHD of ‘sugar sweetened beverages’ in men. To disregard one while upholding the other at the individual food group level is thus logically and methodologically incongruous, and thus interpretations of intact indices must be considered without being able to make inferences about individual items within the index.

Others comparing plant-based dietary indices have also shown variability and inconsistencies. Kim and colleagues (24) contrasted the association of 5 different plant-based dietary indices previously developed by others, namely the 3 indices of the scoring approach (PDI, hPDI, uPDI) (2), provegetarian diet index (1), and overall PDI (i.e., PDI-Rotterdam) (25). They assessed whether these indices are associated with risk of incident hypertension. Participants above the highest vs the lowest quintile PDI, hPDI, and provegetarian diet index were associated with lower risk of hypertension. No significant association was found for the PDI-Rotterdam, and uPDI associations were inconsistent depending on the model. The authors concluded that operational differences could affect indices’ ability to detect diet and disease associations. Still, it remained unclear how one should interpret similar associations detected for differently operationalized indices. “Finding no association between the PDI-Rotterdam and hypertension” was described as ‘surprising’ because PDI-Rotterdam had shown moderate to strong correlations with other indices which had association with hypertension risk, and that in general, there have been strong inverse association between plant-rich dietary patterns that include dairy (e.g., DASH diet) and hypertension risk. If the results obtained from indices should align with pre-existing evidence, it is important to evaluate the information provided by these indices, and whether the inconsistencies stem from methodological limitations or if they offer valuable new insights. If the indices do not carry any better evidence than a random food classification, emergence of inconsistent evidence only results in confusion, which requires researchers to speculate, selectively report, and deviate from objective interpretation of evidence.

One key finding of our agnostic permutations approach is that every scoring method yielded various associations between food groups and CHD, so that each food group can be linked to both beneficial and harmful effects on CHD risk through multiple scores. For instance, there are indices with statistically significant associations with CHD, where one has whole grains positively coded and the other has whole grains reversely coded. Similarly, multiple scores for potatoes show statistically significant associations with CHD in both positive and reverse coding scenarios. The direction of change in associations sometimes aligns with expectations, and sometimes does not.

Our methodological evaluation of a plant-based diet scoring approach revealed that alterations of the scoring approach result in varied and unpredictable diverse associations between food groups and CHD risk. Each food group demonstrated the potential for being both beneficially and deleteriously associated with CHD risk. The observed variability in association directions underscores the potential for ‘guilt by association,’ (or, conversely, ‘benefit by association’) in which any of the food groups we studied could be categorized with either ‘healthy’ or ‘unhealthy’ foods and be found to be part of a dietary index beneficially or deleteriously associated with CHD.

“*If the same mathematical pattern can yield such disparate interpretations, what claim can either have upon reality?*” - Stephen Jay Gould, *Mismeasure of Man* (*26*)

## Acknowledgements

The REGARDS study is supported by cooperative agreement U01 NS041588 co-funded by the National Institute of Neurological Disorders and Stroke (NINDS) and the National Institute on Aging (NIA), National Institutes of Health, Department of Health and Human Service. Additional funding for REGARDS CHD outcomes was provided by R01HL080477. The content is solely the responsibility of the authors and does not necessarily represent the official views of the NINDS, NHLBI, or the NIA. Representatives of the NINDS were involved in the review of the manuscript but were not directly involved in the collection, management, analysis or interpretation of the data. The authors thank the other investigators, the staff, and the participants of the REGARDS study for their valuable contributions. A full list of participating REGARDS investigators and institutions can be found at: https://www.uab.edu/soph/regardsstudy/

AWB, DBA, and YJ-N designed research; BH analyzed data; YJ-N, AWB, and BH wrote the paper; DBA, JMS, AWB, YJ-N, and BH reviewed and edited the manuscript. AWB and YJ-N had primary responsibility for final content. All authors read and approved the final manuscript.

## Data Availability

Researchers who wish to reproduce our analyses can submit a project proposal to the REGARDS team (https://www.uab.edu/soph/regardsstudy/). We did not have any access privilege that others would not have. The analytic code is publicly available at https://osf.io/c3b4z/.

## Funding

The present analysis is supported by the Alliance for Potato Research & Education. The funder did not have any role in design and conduct of the analyses; interpretation of the results; preparation of the manuscript; and decision to submit the manuscript for publication.

## Author Disclosures

In the 36 months prior to the initial submission, DBA has received personal payments or promises for same from: Amin Talati Wasserman for KSF Acquisition Corp (Glanbia); General Mills; Kaleido Biosciences; Law Offices of Ronald Marron; Medpace/Gelesis; Novo Nordisk Foundation; and Zero Longevity Science (as stock options). Donations to a foundation have been made on his behalf by the Northarvest Bean Growers Association. DBA’s institution, Indiana University, and the Indiana University Foundation have received funds or donations to support his research or educational activities from: Alliance for Potato Research and Education; American Egg Board; Arnold Ventures; Eli Lilly and Company; Mars, Inc.; National Cattlemen’s Beef Association; National Pork Board; Pfizer, Inc.; USDA; WW (formerly Weight Watchers); and numerous other for-profit and non-profit organizations to support the work of the School of Public Health and the university more broadly. AWB has received travel expenses from Alliance for Potato Research and Education, International Food Information Council, International Food Information Council Foundation, and Soy Nutrition Institute Global; speaking honoraria from Alliance for Potato Research and Education, American Society for Nutrition, Calorie Control Council, Eastern North American Region of the International Biometric Society, International Food Information Council Foundation, Potatoes USA, Purchaser Business Group on Health, The Obesity Society, and University of Arkansas for Medical Sciences; consulting payments from National Cattlemen’s Beef Association, and Soy Nutrition Institute Global; and grants through his institution from Alliance for Potato Research & Education, American Egg Board, National Cattlemen’s Beef Association, NIH/NHLBI, NIH/NIDDK, NIH/NIGMS, and NSF. He has been involved in research for which his institution or colleagues have received grants or contracts from Alliance for Potato Research & Education, Gordon and Betty Moore Foundation, Hass Avocado Board, NIH/NCATS, NIH/NCI, NIH/NIA, NIH/NIGMS, and NIH/NLM. His wife is employed by Reckitt. BH has been involved in research for which her institution, Indiana University, has received grants from NIH; WW International, Inc., the National Pork Board, the California Walnut Commission, the Alliance of Potato Research & Education, and the National Cattlemen’s Beef Association. YJ-N has received honoraria from The Alliance for Potato Research and Education. JMS reports no disclosures.

## Abbreviations

BMI: Body mass index
CHD: Coronary heart disease
CI: Confidence interval
ECG: Electrocardiogram
hPDI: Healthful plant-based diet index
HPFS: Health Professionals Follow-Up Study
HR: Hazard ratio
MI: Myocardial infarction
NHS: Nurses’ Health Study
NHS2: Nurses’ Health Study 2
PDI: Plant-based diet index
REGARDS: REasons for Geographic and Racial Differences in Stroke
uPDI: Unhealthful plant-based diet index

## Footnotes

1 Satija et al. excluded individuals with cancer but with an exception for nonmelanoma skin cancer, we could not distinguish among cancer types in the REGARDS data we received.

2 REGARDS question: “During the past year, have you taken any vitamins or minerals regularly, at least once a month?” (yes, no).

3 Quintiles were used, but reported intakes below q1 and q2 were combined because of imbalance in the number of participants from zero consumption.

4 Has diabetes if fasting glucose>=126/non-fasting glucose>=200 or pills or insulin, self-reported diabetes and taking insulin, self-reported diabetes and taking glucose lowering pills, or self-reported diabetes.

5 Has dyslipidemia if TC≥240 or LDL≥160 or HDL≤40 or on medication, self-reported elevated lipids, or self-reported use of lipid lowering medication only in those answering yes to having lipidemia.

6 Has hypertension if SBP≥140 or DBP≥90 or self-reported current medication use to control blood pressure.

7 REGARDS dataset includes a variable for ‘[number of] alcoholic drinks per week’. We divided values by 7 (to obtain per day value) and multiplied the result by 14 to obtain gram alcohol consumed per day (National Institute on Alcohol Abuse and Alcoholism. What is a standard drink? https://www.niaaa.nih.gov/alcohols-effects-health/overview-alcohol-consumption/what-standard-drink, Accessed June 21, 2023). We then categorized gram alcohol values per Satija et al.: 0, >0 to <5, 5 to <10, 10 to <15, or ≥15 g/day.

8 Stroke belt (North Carolina, South Carolina, Georgia, Tennessee, Alabama, Mississippi, Arkansas, and Louisiana), stroke buckle (the coastal plain of North Carolina, South Carolina, and Georgia), and from elsewhere in the continental US.

9 REGARDS FFQ item: “White potatoes not fried, incl. boiled, baked, mashed and potato salad”.

10 REGARDS FFQ item: “French fries, fried potatoes or hash browns”.

11 REGARDS FFQ item: “Snack like potato chips, corn chips, popcorn (not pretzels)”.

12 REGARDS FFQ item: “Snack like potato chips, corn chips, popcorn (not pretzels) - low fat”.

## Notes

### Author Declarations

Protocol of the current analysis was approved by Indiana University Institutional Review Board, IRB#16198.

