## Supplementary Tables 1-8 and Supplementary Figures 1-6 for "Guilt by association: Plant-based foods can be incorporated into both healthy and unhealthy plant-based diet indices associated with coronary heart disease"

| **Supplementary Table 1. Median values for each decile^1^ of PDI, hPDI and uPDI by gender and model variation (Recategorization approach)** | | | | | | | | | | | |
| --- | --- | --- | --- | --- | --- | --- | --- | --- | --- | --- | --- |
| **Outcome** | **Model** | **Decile 1** | **Decile 2** | **Decile 3** | **Decile 4** | **Decile 5** | **Decile 6** | **Decile 7** | **Decile 8** | **Decile 9** | **Decile 10** |
| **Women** | | | | | | | | | | | |
| PDI | Replication | 44.0 | 48.0 | 50.0 | 51.0 | 53.0 | 54.0 | 55.5 | 57.0 | 59.0 | 63.0 |
|  | Model 1 | 43.4 | 47.6 | 49.8 | 50.8 | 52.9 | 54.0 | 55.6 | 57.2 | 59.3 | 63.0 |
|  | Model 2 | 44.0 | 48.0 | 50.0 | 51.0 | 53.0 | 54.0 | 55.5 | 57.0 | 59.0 | 63.0 |
|  | Model 3 | 43.4 | 47.6 | 49.8 | 50.8 | 52.9 | 54.0 | 55.6 | 57.2 | 59.3 | 63.0 |
|  | Model 4 | 44.5 | 47.4 | 49.3 | 51.2 | 53.1 | 54.0 | 55.9 | 57.3 | 59.7 | 63.0 |
|  | Model 5 | 44.0 | 48.0 | 50.0 | 51.0 | 53.0 | 54.0 | 56.0 | 57.5 | 59.0 | 63.0 |
| hPDI | Replication | 41.0 | 46.0 | 48.0 | 51.0 | 53.0 | 55.0 | 57.0 | 58.5 | 61.0 | 65.5 |
|  | Model 1 | 41.3 | 46.1 | 48.7 | 50.8 | 52.9 | 54.0 | 56.1 | 58.2 | 61.4 | 65.6 |
|  | Model 2 | 43.0 | 47.0 | 49.0 | 51.0 | 52.5 | 54.0 | 56.0 | 57.5 | 60.0 | 64.0 |
|  | Model 3 | 41.3 | 46.6 | 48.7 | 50.8 | 52.4 | 54.0 | 56.1 | 57.7 | 60.4 | 64.6 |
|  | Model 4 | 42.6 | 46.9 | 49.3 | 51.2 | 53.1 | 54.9 | 56.4 | 58.3 | 60.6 | 64.4 |
|  | Model 5 | 41.0 | 46.0 | 48.0 | 51.0 | 53.0 | 55.0 | 56.5 | 58.5 | 61.0 | 65.5 |
| uPDI | Replication | 40.0 | 45.0 | 48.0 | 50.5 | 52.5 | 54.5 | 56.5 | 59.0 | 61.5 | 66.0 |
|  | Model 1 | 39.2 | 44.5 | 47.6 | 49.8 | 52.4 | 55.1 | 57.2 | 59.3 | 62.5 | 66.7 |
|  | Model 2 | 40.0 | 44.0 | 47.0 | 50.0 | 52.0 | 54.5 | 56.5 | 59.0 | 62.0 | 67.0 |
|  | Model 3 | 39.2 | 44.5 | 47.6 | 49.8 | 52.4 | 54.5 | 57.2 | 59.3 | 62.5 | 66.7 |
|  | Model 4 | 40.7 | 45.0 | 47.8 | 50.2 | 52.1 | 54.5 | 56.4 | 58.7 | 61.6 | 66.3 |
|  | Model 5 | 40.0 | 45.0 | 48.0 | 50.5 | 53.0 | 54.5 | 56.5 | 59.0 | 62.0 | 66.0 |
| **Men** | | | | | | | | | | | |
| PDI | Replication | 44.0 | 48.0 | 50.0 | 51.0 | 53.0 | 54.0 | 56.0 | 57.5 | 59.3 | 63.0 |
|  | Model 1 | 43.4 | 47.6 | 49.8 | 50.8 | 52.9 | 54.0 | 55.6 | 57.2 | 59.3 | 63.0 |
|  | Model 2 | 44.0 | 48.0 | 50.0 | 51.0 | 53.0 | 54.0 | 56.0 | 57.5 | 59.3 | 63.0 |
|  | Model 3 | 43.4 | 47.6 | 49.8 | 50.8 | 52.9 | 54.0 | 55.6 | 57.2 | 59.3 | 63.5 |
|  | Model 4 | 44.5 | 47.4 | 49.3 | 51.2 | 53.1 | 54.0 | 55.9 | 57.3 | 59.7 | 63.5 |
|  | Model 5 | 44.0 | 48.0 | 50.0 | 51.0 | 53.0 | 54.0 | 56.0 | 57.5 | 59.0 | 63.0 |
| hPDI | Replication | 42.0 | 46.0 | 48.0 | 51.0 | 53.0 | 55.0 | 56.8 | 58.5 | 61.0 | 65.0 |
|  | Model 1 | 42.4 | 46.1 | 48.7 | 50.8 | 52.9 | 54.0 | 56.1 | 58.2 | 61.4 | 65.6 |
|  | Model 2 | 43.0 | 47.0 | 49.0 | 51.0 | 52.5 | 54.0 | 56.0 | 57.5 | 60.0 | 64.0 |
|  | Model 3 | 42.4 | 46.6 | 48.7 | 50.8 | 52.9 | 54.0 | 56.1 | 57.7 | 60.4 | 64.6 |
|  | Model 4 | 42.6 | 46.9 | 49.3 | 51.2 | 52.6 | 54.5 | 56.4 | 58.3 | 60.6 | 64.4 |
|  | Model 5 | 41.0 | 46.0 | 48.0 | 50.5 | 53.0 | 54.5 | 56.5 | 58.5 | 61.0 | 65.0 |
| uPDI | Replication | 40.5 | 45.0 | 48.0 | 50.5 | 53.0 | 54.5 | 57.0 | 59.0 | 61.5 | 66.0 |
|  | Model 1 | 40.2 | 44.5 | 47.6 | 49.8 | 52.9 | 55.1 | 57.2 | 59.3 | 62.5 | 66.7 |
|  | Model 2 | 40.0 | 44.0 | 47.0 | 50.0 | 52.5 | 54.5 | 57.0 | 59.0 | 62.0 | 67.0 |
|  | Model 3 | 40.2 | 44.5 | 47.6 | 49.8 | 52.4 | 54.5 | 56.6 | 59.3 | 62.5 | 66.7 |
|  | Model 4 | 40.7 | 44.5 | 47.8 | 50.2 | 52.6 | 54.5 | 56.4 | 58.7 | 61.6 | 66.3 |
|  | Model 5 | 41.0 | 45.0 | 48.0 | 50.5 | 53.0 | 54.5 | 56.5 | 59.0 | 62.0 | 66.0 |
| Notes: Replication: Per the scoring approach advanced by Satija and colleagues. Model 1: Exclude all items in “Potatoes” group from indices. Model 2: Move “Potatoes” group as a whole intact group into “Healthy” super category. Model 3: Move all items in “Potatoes” group into “Vegetables” group. Model 4: Split up “Potatoes” group: move white potato as a single item group into “Healthy” super category, keep French fries and salty snacks in “Potatoes” under “less-healthy” super category. Model 5: Split up “Potatoes” group: move white potato into “Vegetables” group, keep French fries and salty snacks in “Potatoes” under “less-healthy” super category. 1). Note that although decile denotes the 9 values that separate data into 10 equal segments, herein we refer to ‘deciles’ as representing approximately 10% of the data per conventions in the field (that is, ‘decile 1’ represents approximately 10% of observations that fall below decile 1 and ‘decile 10’ represents approximately 10% of observations that fall above decile 9). | | | | | | | | | | | |

**Supplementary Table 2. Multivariable-adjusted^1^ Hazard ratios and 95% confidence interval for CHD according to deciles of the PDI (Recategorization approach)**

| **Model** | **Decile 1** | **Decile 2** | **Decile 3** | **Decile 4** | **Decile 5** | **Decile 6** | **Decile 7** | **Decile 8** | **Decile 9** | **Decile 10** | **HR unit=10** | **P trend** |
| --- | --- | --- | --- | --- | --- | --- | --- | --- | --- | --- | --- | --- |
| **Women** | | | | | | | | | | | | |
| Replication | 1.00 | 0.83 (0.54-1.29) | 0.92 (0.59-1.42) | 0.61 (0.36-1.02) | 0.73 (0.47-1.13) | 0.59 (0.35-1.01) | 0.72 (0.46-1.11) | 0.71 (0.44-1.13) | 0.72 (0.45-1.16) | 0.64 (0.4-1.03) | 0.82 (0.67-1.01) | 0.062 |
| Model 1 | 1.00 | 0.93 (0.6-1.46) | 0.75 (0.47-1.19) | 0.7 (0.43-1.15) | 0.63 (0.4-1.01) | 0.95 (0.59-1.54) | 0.8 (0.52-1.24) | 0.53 (0.31-0.88) | 0.75 (0.46-1.21) | 0.68 (0.42-1.1) | 0.82 (0.67-1) | 0.087 |
| Model 2 | 1.00 | 0.83 (0.54-1.29) | 0.92 (0.59-1.42) | 0.61 (0.36-1.02) | 0.73 (0.47-1.13) | 0.59 (0.35-1.01) | 0.72 (0.46-1.11) | 0.71 (0.44-1.13) | 0.72 (0.45-1.16) | 0.64 (0.4-1.03) | 0.82 (0.67-1.01) | 0.062 |
| Model 3 | 1.00 | 0.81 (0.52-1.27) | 0.75 (0.48-1.17) | 0.65 (0.39-1.06) | 0.66 (0.42-1.05) | 0.72 (0.44-1.19) | 0.82 (0.54-1.25) | 0.48 (0.29-0.82) | 0.82 (0.52-1.3) | 0.61 (0.37-0.99) | 0.82 (0.67-1) | 0.092 |
| Model 4 | 1.00 | 1.15 (0.72-1.84) | 1.09 (0.71-1.65) | 0.91 (0.56-1.49) | 0.86 (0.55-1.34) | 1.02 (0.63-1.62) | 0.69 (0.43-1.12) | 0.89 (0.56-1.43) | 0.83 (0.52-1.33) | 0.83 (0.51-1.35) | 0.82 (0.66-1.02) | 0.129 |
| Model 5 | 1.00 | 0.93 (0.6-1.45) | 1.11 (0.72-1.72) | 0.57 (0.33-0.98) | 0.67 (0.42-1.06) | 0.9 (0.57-1.44) | 0.74 (0.47-1.18) | 0.72 (0.44-1.16) | 0.84 (0.51-1.36) | 0.63 (0.38-1.04) | 0.82 (0.66-1.01) | 0.053 |
| **Men** | | | | | | | | | | | | |
| Replication | 1.00 | 1.25 (0.84-1.86) | 0.87 (0.55-1.37) | 1 (0.63-1.59) | 1.16 (0.77-1.73) | 0.93 (0.58-1.5) | 0.91 (0.6-1.4) | 1.19 (0.77-1.83) | 1.07 (0.69-1.66) | 1.05 (0.69-1.62) | 1.02 (0.85-1.21) | 0.976 |
| Model 1 | 1.00 | 1.02 (0.67-1.56) | 1.08 (0.71-1.63) | 1.13 (0.73-1.76) | 0.71 (0.46-1.1) | 0.98 (0.62-1.57) | 0.96 (0.64-1.45) | 1.07 (0.69-1.64) | 0.94 (0.6-1.46) | 1.03 (0.68-1.57) | 0.99 (0.84-1.17) | 0.972 |
| Model 2 | 1.00 | 1.25 (0.84-1.86) | 0.87 (0.55-1.37) | 1 (0.63-1.59) | 1.16 (0.77-1.73) | 0.93 (0.58-1.5) | 0.91 (0.6-1.4) | 1.19 (0.77-1.83) | 1.07 (0.69-1.66) | 1.05 (0.69-1.62) | 1.02 (0.85-1.21) | 0.976 |
| Model 3 | 1.00 | 0.8 (0.52-1.22) | 1.04 (0.7-1.56) | 0.92 (0.59-1.44) | 0.7 (0.46-1.06) | 0.91 (0.57-1.44) | 0.8 (0.54-1.2) | 0.98 (0.64-1.5) | 0.82 (0.53-1.25) | 0.89 (0.58-1.35) | 0.98 (0.83-1.16) | 0.674 |
| Model 4 | 1.00 | 0.96 (0.62-1.48) | 0.91 (0.62-1.34) | 1.01 (0.65-1.55) | 0.96 (0.64-1.42) | 0.69 (0.42-1.11) | 0.96 (0.64-1.44) | 0.96 (0.63-1.45) | 0.82 (0.54-1.25) | 1.1 (0.73-1.67) | 1.02 (0.85-1.22) | 0.992 |
| Model 5 | 1.00 | 1.01 (0.67-1.52) | 1.2 (0.79-1.81) | 0.72 (0.44-1.19) | 1.08 (0.73-1.61) | 0.9 (0.56-1.42) | 0.77 (0.5-1.2) | 1.19 (0.79-1.81) | 0.77 (0.49-1.24) | 1.11 (0.73-1.69) | 0.99 (0.83-1.18) | 0.958 |
| **Combined** | | | | | | | | | | | | |
| Replication | 1.00 | 1.04 (0.77-1.39) | 0.89 (0.65-1.23) | 0.8 (0.57-1.13) | 0.94 (0.69-1.26) | 0.76 (0.53-1.08) | 0.81 (0.6-1.1) | 0.94 (0.68-1.29) | 0.89 (0.65-1.23) | 0.84 (0.61-1.16) | 0.93 (0.81-1.06) | 0.220 |
| Model 1 | 1.00 | 0.98 (0.72-1.33) | 0.92 (0.67-1.24) | 0.92 (0.66-1.27) | 0.67 (0.49-0.92) | 0.97 (0.69-1.35) | 0.89 (0.66-1.19) | 0.8 (0.57-1.11) | 0.85 (0.61-1.17) | 0.86 (0.63-1.18) | 0.92 (0.81-1.04) | 0.265 |
| Model 2 | 1.00 | 1.04 (0.77-1.39) | 0.89 (0.65-1.23) | 0.8 (0.57-1.13) | 0.94 (0.69-1.26) | 0.76 (0.53-1.08) | 0.81 (0.6-1.1) | 0.94 (0.68-1.29) | 0.89 (0.65-1.23) | 0.84 (0.61-1.16) | 0.93 (0.81-1.06) | 0.220 |
| Model 3 | 1.00 | 0.8 (0.59-1.09) | 0.9 (0.67-1.21) | 0.78 (0.56-1.09) | 0.69 (0.5-0.93) | 0.81 (0.58-1.14) | 0.81 (0.61-1.08) | 0.74 (0.53-1.03) | 0.82 (0.6-1.12) | 0.76 (0.55-1.04) | 0.91 (0.8-1.04) | 0.162 |
| Model 4 | 1.00 | 1.04 (0.76-1.43) | 0.99 (0.74-1.31) | 0.96 (0.7-1.33) | 0.91 (0.68-1.23) | 0.84 (0.6-1.17) | 0.84 (0.62-1.14) | 0.93 (0.68-1.27) | 0.83 (0.6-1.13) | 0.98 (0.71-1.34) | 0.93 (0.81-1.07) | 0.320 |
| Model 5 | 1.00 | 0.97 (0.72-1.31) | 1.16 (0.86-1.56) | 0.65 (0.45-0.94) | 0.88 (0.65-1.2) | 0.9 (0.65-1.25) | 0.76 (0.55-1.04) | 0.96 (0.7-1.32) | 0.8 (0.57-1.12) | 0.88 (0.64-1.22) | 0.92 (0.8-1.05) | 0.196 |

Notes: Replication: Per the classification approach advanced by Satija and colleagues (referred to as the scoring approach in the main text). Model 1: Exclude all items in “Potatoes” group from indices. Model 2: Move “Potatoes” group as a whole intact group into “Healthy” super category. Model 3: Move all items in “Potatoes” group into “Vegetables” group. Model 4: Split up “Potatoes” group: move white potato as a single item group into “Healthy” super category, keep French fries and salty snacks in “Potatoes” under “less-healthy” super category. Model 5: Split up “Potatoes” group: move white potato into “Vegetables” group, keep French fries and salty snacks in “Potatoes” under “less-healthy” super category. 1) Multivariable-adjusted models were adjusted for age (continuous), multivitamin use, family history of MI (yes vs no), margarine intake (in quintiles), diabetes at baseline, hypercholesterolemia at baseline, hypertension at baseline, updated smoking, updated physical activity, updated alcohol intake, updated calorie intake (in quintiles), updated Aspirin user, updated body mass index category, race, updated Regards region. Models for women also adjusted for hormone use and past oral contraceptive use.

**Supplementary Table 3. Multivariable-adjusted^1^ Hazard ratios and 95% confidence interval for CHD according to deciles of the hPDI (Recategorization approach)**

| **Model** | **Decile 1** | **Decile 2** | **Decile 3** | **Decile 4** | **Decile 5** | **Decile 6** | **Decile 7** | **Decile 8** | **Decile 9** | **Decile 10** | **HR unit=10** | **P trend** |
| --- | --- | --- | --- | --- | --- | --- | --- | --- | --- | --- | --- | --- |
| **Women** | | | | | | | | | | | | |
| Replication | 1.00 | 0.9 (0.57-1.42) | 0.86 (0.55-1.33) | 0.62 (0.38-1.03) | 0.74 (0.46-1.17) | 0.73 (0.46-1.16) | 0.59 (0.36-0.97) | 0.62 (0.37-1.04) | 0.64 (0.39-1.04) | 0.82 (0.51-1.33) | 0.9 (0.76-1.07) | 0.172 |
| Model 1 | 1.00 | 0.77 (0.49-1.19) | 0.68 (0.43-1.09) | 0.68 (0.43-1.09) | 0.66 (0.42-1.02) | 0.81 (0.5-1.32) | 0.69 (0.44-1.09) | 0.54 (0.34-0.85) | 0.56 (0.35-0.91) | 0.81 (0.51-1.29) | 0.9 (0.76-1.06) | 0.210 |
| Model 2 | 1.00 | 0.98 (0.63-1.51) | 0.6 (0.36-0.99) | 0.85 (0.53-1.35) | 0.81 (0.52-1.26) | 0.78 (0.47-1.28) | 0.68 (0.42-1.1) | 0.8 (0.51-1.27) | 0.59 (0.37-0.95) | 0.88 (0.56-1.38) | 0.89 (0.74-1.07) | 0.282 |
| Model 3 | 1.00 | 0.88 (0.57-1.36) | 0.61 (0.38-1) | 0.74 (0.46-1.17) | 0.69 (0.44-1.08) | 0.79 (0.49-1.3) | 0.7 (0.44-1.11) | 0.63 (0.39-1.03) | 0.55 (0.34-0.88) | 0.8 (0.51-1.25) | 0.9 (0.75-1.06) | 0.200 |
| Model 4 | 1.00 | 0.94 (0.62-1.44) | 0.66 (0.41-1.07) | 0.59 (0.36-0.96) | 0.91 (0.6-1.39) | 0.61 (0.39-0.95) | 0.7 (0.44-1.11) | 0.67 (0.42-1.07) | 0.58 (0.36-0.93) | 0.84 (0.54-1.31) | 0.89 (0.74-1.06) | 0.249 |
| Model 5 | 1.00 | 1.04 (0.67-1.63) | 0.66 (0.41-1.06) | 0.75 (0.46-1.21) | 0.75 (0.47-1.19) | 0.61 (0.37-0.99) | 0.71 (0.44-1.16) | 0.59 (0.36-0.99) | 0.69 (0.43-1.11) | 0.74 (0.45-1.19) | 0.89 (0.75-1.06) | 0.088 |
| **Men** | | | | | | | | | | | | |
| Replication | 1.00 | 1.03 (0.68-1.57) | 0.83 (0.55-1.26) | 0.93 (0.61-1.41) | 0.67 (0.42-1.06) | 0.84 (0.56-1.28) | 0.8 (0.52-1.24) | 0.9 (0.57-1.4) | 0.61 (0.38-0.97) | 0.8 (0.51-1.26) | 0.89 (0.76-1.03) | 0.089 |
| Model 1 | 1.00 | 0.98 (0.65-1.47) | 0.87 (0.56-1.35) | 0.89 (0.58-1.37) | 0.99 (0.66-1.48) | 0.82 (0.51-1.3) | 0.92 (0.61-1.39) | 0.76 (0.49-1.15) | 0.88 (0.57-1.35) | 0.85 (0.54-1.34) | 0.9 (0.78-1.05) | 0.321 |
| Model 2 | 1.00 | 0.96 (0.63-1.46) | 0.82 (0.53-1.27) | 0.88 (0.57-1.36) | 0.87 (0.58-1.32) | 0.88 (0.55-1.39) | 0.85 (0.56-1.3) | 0.71 (0.45-1.11) | 0.91 (0.6-1.39) | 0.81 (0.53-1.25) | 0.91 (0.78-1.07) | 0.313 |
| Model 3 | 1.00 | 0.93 (0.62-1.4) | 0.77 (0.49-1.2) | 0.8 (0.52-1.24) | 0.91 (0.61-1.36) | 0.77 (0.48-1.22) | 0.78 (0.51-1.2) | 0.74 (0.48-1.15) | 0.85 (0.56-1.29) | 0.73 (0.47-1.12) | 0.89 (0.77-1.04) | 0.187 |
| Model 4 | 1.00 | 1.01 (0.67-1.52) | 0.87 (0.56-1.36) | 1.12 (0.74-1.71) | 0.95 (0.63-1.44) | 0.88 (0.58-1.35) | 0.84 (0.55-1.29) | 0.89 (0.57-1.38) | 0.93 (0.6-1.43) | 0.87 (0.56-1.35) | 0.91 (0.77-1.06) | 0.375 |
| Model 5 | 1.00 | 0.95 (0.6-1.48) | 0.96 (0.63-1.47) | 1.04 (0.67-1.6) | 0.9 (0.58-1.4) | 0.93 (0.6-1.43) | 0.69 (0.43-1.11) | 0.8 (0.5-1.26) | 0.96 (0.61-1.51) | 0.81 (0.51-1.29) | 0.89 (0.77-1.04) | 0.192 |
| **Combined** | | | | | | | | | | | | |
| Replication | 1.00 | 0.97 (0.71-1.32) | 0.84 (0.62-1.14) | 0.79 (0.57-1.09) | 0.7 (0.51-0.97) | 0.79 (0.58-1.08) | 0.7 (0.51-0.97) | 0.77 (0.55-1.08) | 0.62 (0.44-0.87) | 0.81 (0.58-1.13) | 0.89 (0.8-1) | 0.029 |
| Model 1 | 1.00 | 0.87 (0.64-1.18) | 0.78 (0.57-1.07) | 0.79 (0.57-1.08) | 0.82 (0.61-1.1) | 0.82 (0.58-1.14) | 0.81 (0.59-1.09) | 0.65 (0.48-0.88) | 0.72 (0.52-0.99) | 0.83 (0.6-1.15) | 0.9 (0.8-1.01) | 0.116 |
| Model 2 | 1.00 | 0.97 (0.71-1.31) | 0.72 (0.52-1) | 0.87 (0.63-1.19) | 0.84 (0.62-1.14) | 0.83 (0.59-1.17) | 0.77 (0.56-1.06) | 0.75 (0.54-1.04) | 0.75 (0.55-1.03) | 0.84 (0.62-1.15) | 0.9 (0.8-1.02) | 0.142 |
| Model 3 | 1.00 | 0.9 (0.67-1.22) | 0.69 (0.5-0.96) | 0.77 (0.56-1.06) | 0.8 (0.59-1.08) | 0.78 (0.56-1.09) | 0.74 (0.54-1.02) | 0.69 (0.5-0.95) | 0.7 (0.51-0.95) | 0.76 (0.56-1.04) | 0.89 (0.8-1) | 0.066 |
| Model 4 | 1.00 | 0.98 (0.73-1.31) | 0.77 (0.56-1.07) | 0.85 (0.62-1.17) | 0.93 (0.69-1.25) | 0.74 (0.55-1.01) | 0.77 (0.56-1.06) | 0.78 (0.56-1.08) | 0.75 (0.54-1.03) | 0.85 (0.62-1.17) | 0.9 (0.8-1.01) | 0.153 |
| Model 5 | 1.00 | 0.99 (0.72-1.36) | 0.81 (0.59-1.12) | 0.89 (0.65-1.23) | 0.82 (0.6-1.13) | 0.77 (0.56-1.07) | 0.7 (0.5-0.99) | 0.7 (0.5-0.99) | 0.82 (0.59-1.14) | 0.77 (0.55-1.08) | 0.89 (0.8-1) | 0.035 |

Notes: Replication: Per the classification approach advanced by Satija and colleagues (referred to as the scoring approach in the main text). Model 1: Exclude all items in “Potatoes” group from indices. Model 2: Move “Potatoes” group as a whole intact group into “Healthy” super category. Model 3: Move all items in “Potatoes” group into “Vegetables” group. Model 4: Split up “Potatoes” group: move white potato as a single item group into “Healthy” super category, keep French fries and salty snacks in “Potatoes” under “less-healthy” super category. Model 5: Split up “Potatoes” group: move white potato into “Vegetables” group, keep French fries and salty snacks in “Potatoes” under “less-healthy” super category. 1) Multivariable-adjusted models were adjusted for age (continuous), multivitamin use, family history of MI (yes vs no), margarine intake (in quintiles), diabetes at baseline, hypercholesterolemia at baseline, hypertension at baseline, updated smoking, updated physical activity, updated alcohol intake, updated calorie intake (in quintiles), updated Aspirin user, updated body mass index category, race, updated Regards region. Models for women also adjusted for hormone use and past oral contraceptive use.

**Supplementary Table 4. Multivariable-adjusted^1^ Hazard ratios and 95% confidence intervals for CHD according to deciles of the uPDI (Recategorization approach)**

| **Model** | **Decile 1** | **Decile 2** | **Decile 3** | **Decile 4** | **Decile 5** | **Decile 6** | **Decile 7** | **Decile 8** | **Decile 9** | **Decile 10** | **HR unit=10** | **P trend** |
| --- | --- | --- | --- | --- | --- | --- | --- | --- | --- | --- | --- | --- |
| **Women** | | | | | | | | | | | | |
| Replication | 1.00 | 1.24 (0.74-2.07) | 1.21 (0.73-2.02) | 1.14 (0.67-1.94) | 1.15 (0.68-1.95) | 0.94 (0.54-1.62) | 1.31 (0.78-2.22) | 1.41 (0.84-2.37) | 1.27 (0.74-2.16) | 1.41 (0.83-2.38) | 1.1 (0.94-1.29) | 0.198 |
| Model 1 | 1.00 | 0.62 (0.37-1.02) | 0.92 (0.56-1.5) | 0.91 (0.58-1.42) | 0.71 (0.43-1.18) | 0.86 (0.53-1.39) | 0.92 (0.55-1.52) | 1.01 (0.63-1.61) | 0.92 (0.56-1.51) | 1.02 (0.62-1.67) | 1.1 (0.94-1.28) | 0.347 |
| Model 2 | 1.00 | 0.54 (0.31-0.95) | 0.79 (0.49-1.27) | 0.88 (0.56-1.38) | 0.79 (0.48-1.3) | 0.89 (0.55-1.43) | 1.03 (0.63-1.66) | 0.89 (0.55-1.45) | 0.95 (0.58-1.54) | 0.99 (0.6-1.64) | 1.1 (0.94-1.29) | 0.279 |
| Model 3 | 1.00 | 0.63 (0.38-1.03) | 0.79 (0.48-1.32) | 0.88 (0.56-1.37) | 0.75 (0.45-1.23) | 0.85 (0.52-1.36) | 0.74 (0.44-1.23) | 0.96 (0.6-1.55) | 1.03 (0.63-1.68) | 0.92 (0.56-1.52) | 1.1 (0.94-1.29) | 0.373 |
| Model 4 | 1.00 | 0.81 (0.48-1.36) | 1.09 (0.68-1.74) | 0.86 (0.53-1.4) | 1.29 (0.81-2.05) | 1.01 (0.62-1.64) | 1.05 (0.63-1.74) | 1.22 (0.76-1.97) | 1.12 (0.69-1.83) | 1.22 (0.74-2.01) | 1.11 (0.95-1.3) | 0.179 |
| Model 5 | 1.00 | 1.04 (0.64-1.7) | 1 (0.61-1.63) | 1.06 (0.63-1.76) | 1.07 (0.66-1.75) | 0.98 (0.58-1.64) | 0.92 (0.54-1.55) | 1.5 (0.93-2.44) | 1.09 (0.66-1.82) | 1.21 (0.73-2) | 1.11 (0.95-1.29) | 0.275 |
| **Men** | | | | | | | | | | | | |
| Replication | 1.00 | 0.85 (0.54-1.33) | 0.93 (0.6-1.44) | 0.88 (0.55-1.4) | 1.07 (0.69-1.65) | 0.87 (0.55-1.37) | 1.02 (0.65-1.6) | 1.16 (0.75-1.8) | 1.12 (0.71-1.78) | 1.05 (0.66-1.68) | 1.12 (0.97-1.3) | 0.286 |
| Model 1 | 1.00 | 0.94 (0.6-1.48) | 1.26 (0.79-2.01) | 1.06 (0.7-1.62) | 0.95 (0.59-1.52) | 1.14 (0.73-1.78) | 1.2 (0.76-1.9) | 1.19 (0.76-1.86) | 1.32 (0.83-2.09) | 1.12 (0.7-1.82) | 1.1 (0.96-1.26) | 0.316 |
| Model 2 | 1.00 | 1.15 (0.73-1.81) | 1.37 (0.89-2.13) | 1.01 (0.65-1.56) | 1.09 (0.69-1.74) | 1.01 (0.63-1.62) | 1.45 (0.92-2.27) | 1.21 (0.77-1.91) | 1.28 (0.81-2.05) | 1.17 (0.71-1.91) | 1.08 (0.94-1.25) | 0.504 |
| Model 3 | 1.00 | 0.99 (0.63-1.56) | 1.33 (0.83-2.13) | 1.06 (0.68-1.63) | 1.06 (0.65-1.7) | 1.13 (0.71-1.79) | 1.28 (0.81-2.04) | 1.26 (0.8-1.98) | 1.41 (0.88-2.27) | 1.19 (0.73-1.94) | 1.11 (0.96-1.27) | 0.213 |
| Model 4 | 1.00 | 1.05 (0.67-1.66) | 1.26 (0.82-1.93) | 1.17 (0.77-1.77) | 1.04 (0.65-1.66) | 1.14 (0.73-1.78) | 1.15 (0.73-1.81) | 1.11 (0.7-1.75) | 1.37 (0.88-2.14) | 1.21 (0.75-1.95) | 1.09 (0.94-1.26) | 0.351 |
| Model 5 | 1.00 | 1 (0.64-1.57) | 1.19 (0.78-1.83) | 1.08 (0.68-1.69) | 1.24 (0.79-1.93) | 0.96 (0.6-1.53) | 1.13 (0.72-1.78) | 1.07 (0.67-1.71) | 1.31 (0.83-2.06) | 1.26 (0.79-2.01) | 1.11 (0.96-1.28) | 0.275 |
| **Combined** | | | | | | | | | | | | |
| Replication | 1.00 | 1 (0.71-1.4) | 1.04 (0.75-1.45) | 0.98 (0.69-1.4) | 1.1 (0.79-1.54) | 0.9 (0.63-1.27) | 1.14 (0.81-1.6) | 1.26 (0.9-1.76) | 1.18 (0.84-1.68) | 1.19 (0.84-1.69) | 1.11 (1-1.24) | 0.098 |
| Model 1 | 1.00 | 0.78 (0.56-1.09) | 1.08 (0.77-1.52) | 0.99 (0.73-1.34) | 0.83 (0.59-1.17) | 1 (0.72-1.38) | 1.06 (0.76-1.49) | 1.1 (0.79-1.52) | 1.11 (0.8-1.56) | 1.07 (0.76-1.51) | 1.1 (0.99-1.22) | 0.169 |
| Model 2 | 1.00 | 0.85 (0.6-1.21) | 1.06 (0.77-1.47) | 0.95 (0.69-1.29) | 0.94 (0.67-1.32) | 0.95 (0.68-1.33) | 1.23 (0.89-1.71) | 1.05 (0.76-1.46) | 1.11 (0.79-1.55) | 1.08 (0.76-1.53) | 1.09 (0.98-1.21) | 0.223 |
| Model 3 | 1.00 | 0.81 (0.58-1.12) | 1.05 (0.74-1.48) | 0.96 (0.71-1.32) | 0.89 (0.63-1.26) | 0.98 (0.7-1.37) | 1 (0.71-1.41) | 1.11 (0.8-1.54) | 1.21 (0.86-1.7) | 1.05 (0.74-1.49) | 1.1 (1-1.22) | 0.127 |
| Model 4 | 1.00 | 0.94 (0.67-1.32) | 1.18 (0.86-1.61) | 1.02 (0.75-1.41) | 1.16 (0.83-1.61) | 1.08 (0.78-1.5) | 1.1 (0.79-1.55) | 1.16 (0.83-1.62) | 1.25 (0.9-1.74) | 1.21 (0.86-1.71) | 1.1 (0.99-1.23) | 0.111 |
| Model 5 | 1.00 | 1.02 (0.73-1.42) | 1.1 (0.8-1.52) | 1.07 (0.76-1.5) | 1.16 (0.83-1.61) | 0.97 (0.68-1.37) | 1.03 (0.73-1.46) | 1.26 (0.9-1.77) | 1.21 (0.86-1.7) | 1.23 (0.88-1.74) | 1.11 (1-1.23) | 0.123 |

Notes: Replication: Per the classification approach advanced by Satija and colleagues (referred to as the scoring approach in the main text). Model 1: Exclude all items in “Potatoes” group from indices. Model 2: Move “Potatoes” group as a whole intact group into “Healthy” super category. Model 3: Move all items in “Potatoes” group into “Vegetables” group. Model 4: Split up “Potatoes” group: move white potato as a single item group into “Healthy” super category, keep French fries and salty snacks in “Potatoes” under “less-healthy” super category. Model 5: Split up “Potatoes” group: move white potato into “Vegetables” group, keep French fries and salty snacks in “Potatoes” under “less-healthy” super category. 1) Multivariable-adjusted models were adjusted for age (continuous), multivitamin use, family history of MI (yes vs no), margarine intake (in quintiles), diabetes at baseline, hypercholesterolemia at baseline, hypertension at baseline, updated smoking, updated physical activity, updated alcohol intake, updated calorie intake (in quintiles), updated Aspirin user, updated body mass index category, race, updated Regards region. Models for women also adjusted for hormone use and past oral contraceptive use.

**Supplementary Table 5. Median values for each decile^1^ of PDI, hPDI and uPDI by gender and model variation (Leave-one-out approach)**

| **Outcome** | **Excluded food group** | **Decile 1** | **Decile 2** | **Decile 3** | **Decile 4** | **Decile 5** | **Decile 6** | **Decile 7** | **Decile 8** | **Decile 9** | **Decile 10** |
| --- | --- | --- | --- | --- | --- | --- | --- | --- | --- | --- | --- |
| **Women** | | | | | | | | | | | |
| PDI | Replication | 44.0 | 48.0 | 50.0 | 51.0 | 53.0 | 54.0 | 55.5 | 57.0 | 59.0 | 63.0 |
|  | Whole grains | 43.4 | 47.6 | 49.8 | 50.8 | 52.9 | 54.0 | 55.1 | 57.2 | 59.3 | 62.5 |
|  | Fruits | 44.5 | 47.6 | 49.8 | 50.8 | 52.9 | 54.0 | 55.6 | 57.2 | 59.3 | 62.5 |
|  | Vegetables | 43.4 | 47.6 | 49.8 | 50.8 | 52.9 | 54.0 | 55.6 | 57.2 | 59.3 | 62.5 |
|  | Nuts | 43.4 | 47.6 | 49.8 | 50.8 | 52.9 | 54.0 | 55.6 | 57.2 | 59.3 | 62.5 |
|  | Legumes | 44.5 | 47.6 | 49.8 | 50.8 | 52.9 | 54.0 | 55.6 | 57.2 | 59.3 | 62.5 |
|  | Vegetable oils | 43.4 | 47.6 | 49.8 | 50.8 | 52.9 | 54.0 | 55.6 | 57.2 | 59.3 | 63.0 |
|  | Tea & coffee | 43.4 | 47.6 | 49.8 | 50.8 | 52.9 | 54.0 | 55.6 | 57.2 | 59.3 | 63.5 |
|  | Fruit juices | 43.4 | 47.6 | 49.8 | 50.8 | 52.9 | 54.0 | 55.6 | 57.2 | 59.3 | 63.0 |
|  | Refined grains | 43.4 | 47.6 | 49.8 | 50.8 | 52.9 | 54.0 | 55.6 | 57.2 | 59.3 | 63.0 |
|  | Potatoes | 43.4 | 47.6 | 49.8 | 50.8 | 52.9 | 54.0 | 55.6 | 57.2 | 59.3 | 63.0 |
|  | Sugar sw. beverage | 43.4 | 47.6 | 49.8 | 50.8 | 52.9 | 54.0 | 55.6 | 57.2 | 59.3 | 63.5 |
|  | Sweets & desserts | 43.4 | 47.6 | 49.8 | 50.8 | 52.9 | 54.0 | 55.6 | 57.2 | 59.3 | 63.0 |
| hPDI | Replication | 41.0 | 46.0 | 48.0 | 51.0 | 53.0 | 55.0 | 57.0 | 58.5 | 61.0 | 65.5 |
|  | Whole grains | 40.8 | 45.5 | 48.7 | 50.8 | 52.9 | 54.5 | 56.6 | 58.2 | 61.4 | 65.6 |
|  | Fruits | 41.3 | 45.5 | 48.7 | 50.8 | 52.9 | 55.1 | 56.6 | 58.8 | 61.4 | 65.6 |
|  | Vegetables | 41.3 | 45.5 | 48.7 | 50.8 | 52.9 | 55.1 | 56.6 | 58.8 | 61.4 | 65.6 |
|  | Nuts | 40.2 | 45.5 | 48.7 | 50.8 | 52.9 | 55.1 | 56.6 | 58.8 | 61.4 | 65.6 |
|  | Legumes | 41.3 | 45.5 | 48.2 | 50.8 | 52.9 | 54.5 | 56.6 | 58.8 | 61.4 | 65.6 |
|  | Vegetable oils | 40.2 | 45.5 | 48.2 | 50.8 | 52.9 | 54.5 | 56.6 | 58.8 | 61.4 | 65.6 |
|  | Tea & coffee | 41.3 | 45.5 | 47.6 | 50.8 | 52.9 | 55.1 | 56.6 | 58.8 | 61.4 | 66.2 |
|  | Fruit juices | 41.3 | 45.5 | 48.7 | 50.8 | 52.4 | 54.5 | 56.6 | 58.8 | 61.4 | 65.6 |
|  | Refined grains | 41.8 | 46.6 | 48.7 | 50.8 | 52.9 | 54.0 | 56.1 | 58.2 | 60.4 | 65.6 |
|  | Potatoes | 41.3 | 46.1 | 48.7 | 50.8 | 52.9 | 54.0 | 56.1 | 58.2 | 61.4 | 65.6 |
|  | Sugar sw. beverage | 41.3 | 46.1 | 48.7 | 50.8 | 52.9 | 54.0 | 56.1 | 58.2 | 61.4 | 65.6 |
|  | Sweets & desserts | 42.4 | 46.6 | 48.7 | 50.8 | 52.9 | 54.0 | 56.1 | 58.2 | 60.4 | 65.6 |
| uPDI | Replication | 40.0 | 45.0 | 48.0 | 50.5 | 52.5 | 54.5 | 56.5 | 59.0 | 61.5 | 66.0 |
|  | Whole grains | 40.2 | 45.0 | 47.6 | 50.8 | 52.9 | 54.5 | 56.6 | 58.8 | 61.4 | 65.6 |
|  | Fruits | 40.2 | 45.5 | 47.6 | 50.8 | 52.9 | 54.5 | 56.1 | 58.8 | 61.4 | 65.6 |
|  | Vegetables | 41.3 | 45.5 | 48.7 | 50.8 | 52.9 | 54.5 | 56.1 | 58.2 | 61.4 | 65.6 |
|  | Nuts | 40.2 | 45.0 | 47.6 | 50.8 | 52.9 | 55.1 | 56.6 | 58.8 | 61.4 | 65.6 |
|  | Legumes | 40.2 | 45.5 | 47.6 | 50.8 | 52.9 | 54.5 | 56.6 | 58.2 | 61.4 | 65.6 |
|  | Vegetable oils | 40.2 | 45.0 | 47.6 | 50.8 | 52.9 | 54.5 | 56.6 | 58.8 | 61.4 | 66.7 |
|  | Tea & coffee | 40.2 | 45.0 | 47.6 | 50.8 | 52.9 | 54.5 | 56.1 | 58.8 | 61.4 | 66.7 |
|  | Fruit juices | 40.2 | 44.5 | 47.6 | 50.8 | 52.4 | 54.5 | 56.6 | 59.3 | 61.9 | 66.7 |
|  | Refined grains | 39.2 | 44.5 | 47.6 | 49.8 | 52.9 | 54.5 | 56.6 | 59.3 | 62.5 | 66.7 |
|  | Potatoes | 39.2 | 44.5 | 47.6 | 49.8 | 52.4 | 55.1 | 57.2 | 59.3 | 62.5 | 66.7 |
|  | Sugar sw. beverage | 40.2 | 45.0 | 47.6 | 50.8 | 52.9 | 54.5 | 56.6 | 58.8 | 61.4 | 66.7 |
|  | Sweets & desserts | 39.2 | 44.5 | 47.6 | 50.3 | 52.9 | 55.1 | 56.6 | 59.3 | 62.5 | 66.7 |
| **Men** | | | | | | | | | | | |
| PDI | Replication | 44.0 | 48.0 | 50.0 | 51.0 | 53.0 | 54.0 | 56.0 | 57.5 | 59.3 | 63.0 |
|  | Whole grains | 43.9 | 47.6 | 49.8 | 50.8 | 52.9 | 54.0 | 55.6 | 57.2 | 59.3 | 62.5 |
|  | Fruits | 44.5 | 47.6 | 49.8 | 50.8 | 52.9 | 54.0 | 55.6 | 57.2 | 59.3 | 62.5 |
|  | Vegetables | 43.4 | 47.6 | 49.8 | 50.8 | 52.9 | 54.0 | 55.6 | 57.2 | 59.3 | 62.5 |
|  | Nuts | 43.4 | 47.6 | 49.8 | 50.8 | 52.9 | 54.0 | 55.6 | 57.2 | 59.3 | 63.0 |
|  | Legumes | 44.5 | 47.6 | 49.8 | 50.8 | 52.9 | 54.0 | 55.6 | 57.2 | 59.3 | 62.5 |
|  | Vegetable oils | 43.4 | 47.6 | 49.8 | 50.8 | 52.9 | 54.0 | 55.6 | 57.2 | 59.3 | 63.0 |
|  | Tea & coffee | 43.4 | 47.1 | 49.8 | 50.8 | 52.9 | 54.0 | 55.1 | 57.2 | 59.3 | 63.5 |
|  | Fruit juices | 43.4 | 47.6 | 49.8 | 50.8 | 52.9 | 54.0 | 55.6 | 57.2 | 59.3 | 63.5 |
|  | Refined grains | 43.4 | 47.6 | 49.8 | 50.8 | 52.9 | 54.0 | 55.6 | 57.2 | 59.3 | 63.0 |
|  | Potatoes | 43.4 | 47.6 | 49.8 | 50.8 | 52.9 | 54.0 | 55.6 | 57.2 | 59.3 | 63.0 |
|  | Sugar sw. beverage | 43.4 | 47.6 | 49.8 | 50.8 | 52.9 | 54.0 | 55.6 | 57.7 | 59.3 | 63.5 |
|  | Sweets & desserts | 43.4 | 47.6 | 49.8 | 50.8 | 52.9 | 54.0 | 55.6 | 57.2 | 59.3 | 63.5 |
| hPDI | Replication | 42.0 | 46.0 | 48.0 | 51.0 | 53.0 | 55.0 | 56.8 | 58.5 | 61.0 | 65.0 |
|  | Whole grains | 41.3 | 45.5 | 48.7 | 50.8 | 52.9 | 55.1 | 56.6 | 58.8 | 61.4 | 65.6 |
|  | Fruits | 41.3 | 45.5 | 48.7 | 50.8 | 52.4 | 54.5 | 56.6 | 58.8 | 61.4 | 65.6 |
|  | Vegetables | 41.3 | 45.5 | 48.7 | 50.8 | 52.4 | 54.5 | 56.6 | 58.2 | 61.4 | 65.6 |
|  | Nuts | 40.8 | 45.5 | 48.2 | 50.8 | 52.4 | 54.5 | 56.9 | 59.3 | 61.4 | 65.6 |
|  | Legumes | 41.3 | 45.5 | 48.2 | 50.8 | 52.9 | 55.1 | 56.6 | 58.8 | 61.4 | 65.6 |
|  | Vegetable oils | 41.3 | 45.5 | 48.2 | 50.8 | 52.4 | 54.8 | 56.6 | 58.8 | 61.4 | 65.6 |
|  | Tea & coffee | 41.3 | 45.5 | 47.6 | 50.8 | 52.9 | 55.1 | 56.6 | 58.8 | 61.4 | 65.6 |
|  | Fruit juices | 41.6 | 45.5 | 48.7 | 50.8 | 52.9 | 55.1 | 56.6 | 58.2 | 61.4 | 65.6 |
|  | Refined grains | 41.8 | 46.6 | 48.7 | 50.8 | 52.9 | 54.0 | 56.1 | 58.2 | 60.4 | 65.6 |
|  | Potatoes | 42.4 | 46.1 | 48.7 | 50.8 | 52.9 | 54.0 | 56.1 | 58.2 | 61.4 | 65.6 |
|  | Sugar sw. beverage | 42.4 | 46.6 | 48.7 | 50.8 | 52.9 | 54.0 | 56.1 | 58.2 | 61.4 | 65.6 |
|  | Sweets & desserts | 42.4 | 46.6 | 48.7 | 50.8 | 52.9 | 54.0 | 56.1 | 58.2 | 60.4 | 65.6 |
| uPDI | Replication | 40.5 | 45.0 | 48.0 | 50.5 | 53.0 | 54.5 | 57.0 | 59.0 | 61.5 | 66.0 |
|  | Whole grains | 40.2 | 45.0 | 47.6 | 50.8 | 52.9 | 55.1 | 56.6 | 58.2 | 61.4 | 65.6 |
|  | Fruits | 40.8 | 45.5 | 47.6 | 50.8 | 52.4 | 54.5 | 56.1 | 59.3 | 61.4 | 65.6 |
|  | Vegetables | 41.3 | 45.5 | 48.2 | 50.8 | 52.9 | 54.5 | 56.6 | 58.8 | 61.4 | 65.6 |
|  | Nuts | 41.3 | 45.0 | 47.6 | 50.8 | 52.9 | 54.5 | 56.6 | 58.2 | 61.4 | 65.6 |
|  | Legumes | 40.8 | 45.5 | 47.6 | 50.8 | 52.9 | 54.5 | 56.1 | 58.8 | 61.4 | 65.6 |
|  | Vegetable oils | 40.2 | 45.0 | 47.6 | 50.8 | 52.4 | 54.5 | 56.6 | 58.8 | 61.4 | 66.7 |
|  | Tea & coffee | 40.2 | 44.5 | 47.6 | 50.8 | 52.9 | 54.5 | 56.6 | 58.8 | 61.4 | 66.7 |
|  | Fruit juices | 40.2 | 44.5 | 47.6 | 50.3 | 52.9 | 55.1 | 56.6 | 59.3 | 62.5 | 66.7 |
|  | Refined grains | 40.2 | 44.5 | 47.6 | 49.8 | 52.7 | 54.5 | 56.6 | 59.3 | 62.5 | 66.7 |
|  | Potatoes | 40.2 | 44.5 | 47.6 | 49.8 | 52.9 | 55.1 | 57.2 | 59.3 | 62.5 | 66.7 |
|  | Sugar sw. beverage | 40.8 | 44.5 | 47.6 | 50.8 | 52.9 | 54.5 | 56.6 | 58.8 | 61.4 | 65.6 |
|  | Sweets & desserts | 40.2 | 44.5 | 47.6 | 50.3 | 52.9 | 54.5 | 56.6 | 59.3 | 62.5 | 66.7 |
| Notes: Replication: Per the scoring approach advanced by Satija and colleagues. 1) Note that although decile denotes the 9 values that separate data into 10 equal segments, herein we refer to ‘deciles’ as representing approximately 10% of the data per conventions in the field (that is, ‘decile 1’ represents approximately 10% of observations that fall below decile 1 and ‘decile 10’ represents approximately 10% of observations that fall above decile 9). | | | | | | | | | | | |

**Supplementary Table 6. Multivariable-adjusted^1^ Hazard ratios and 95% confidence interval for CHD according to deciles of the PDI (Leave-one-out approach)**

| **Excluded food group** | **Decile 1** | **Decile 2** | **Decile 3** | **Decile 4** | **Decile 5** | **Decile 6** | **Decile 7** | **Decile 8** | **Decile 9** | **Decile 10** | **HR unit=10** | **P trend** |
| --- | --- | --- | --- | --- | --- | --- | --- | --- | --- | --- | --- | --- |
| **Women** | | | | | | | | | | | | |
| Replication | 1.00 | 0.83 (0.54-1.29) | 0.92 (0.59-1.42) | 0.61 (0.36-1.02) | 0.73 (0.47-1.13) | 0.59 (0.35-1.01) | 0.72 (0.46-1.11) | 0.71 (0.44-1.13) | 0.72 (0.45-1.16) | 0.64 (0.4-1.03) | 0.82 (0.67-1.01) | 0.062 |
| Whole grains | 1.00 | 0.7 (0.43-1.13) | 0.87 (0.55-1.36) | 1 (0.62-1.6) | 0.69 (0.44-1.08) | 0.71 (0.42-1.19) | 0.89 (0.58-1.37) | 0.68 (0.39-1.16) | 0.66 (0.41-1.06) | 0.79 (0.49-1.28) | 0.87 (0.71-1.07) | 0.301 |
| Fruits | 1.00 | 1.01 (0.64-1.59) | 0.98 (0.63-1.53) | 0.86 (0.53-1.39) | 0.67 (0.43-1.05) | 0.93 (0.58-1.5) | 0.88 (0.58-1.35) | 0.73 (0.44-1.21) | 0.79 (0.5-1.25) | 0.68 (0.41-1.11) | 0.83 (0.67-1.02) | 0.070 |
| Vegetables | 1.00 | 0.91 (0.57-1.45) | 0.86 (0.55-1.34) | 0.86 (0.52-1.4) | 0.63 (0.4-1.01) | 0.72 (0.43-1.2) | 0.77 (0.49-1.2) | 0.83 (0.52-1.31) | 0.67 (0.41-1.1) | 0.57 (0.35-0.95) | 0.8 (0.65-0.98) | 0.029 |
| Nuts | 1.00 | 0.74 (0.45-1.19) | 0.83 (0.53-1.32) | 1.11 (0.7-1.78) | 0.69 (0.44-1.1) | 0.77 (0.46-1.27) | 0.78 (0.5-1.23) | 0.68 (0.42-1.12) | 0.7 (0.43-1.15) | 0.73 (0.45-1.19) | 0.86 (0.7-1.05) | 0.141 |
| Legumes | 1.00 | 1.04 (0.66-1.63) | 0.85 (0.55-1.34) | 0.71 (0.43-1.17) | 0.83 (0.54-1.27) | 0.84 (0.52-1.36) | 0.77 (0.5-1.2) | 0.71 (0.43-1.2) | 0.81 (0.52-1.27) | 0.65 (0.4-1.05) | 0.81 (0.66-1) | 0.057 |
| Vegetables oils | 1.00 | 0.99 (0.63-1.55) | 0.62 (0.37-1.02) | 0.85 (0.51-1.43) | 0.89 (0.56-1.39) | 0.82 (0.5-1.34) | 0.76 (0.48-1.19) | 0.86 (0.53-1.38) | 0.71 (0.43-1.18) | 0.72 (0.44-1.18) | 0.87 (0.71-1.07) | 0.216 |
| Tea & coffee | 1.00 | 0.61 (0.38-0.98) | 0.85 (0.56-1.29) | 0.68 (0.42-1.12) | 0.76 (0.5-1.18) | 0.71 (0.43-1.16) | 0.67 (0.44-1.03) | 0.6 (0.36-0.98) | 0.67 (0.42-1.07) | 0.61 (0.38-0.99) | 0.83 (0.68-1.01) | 0.080 |
| Fruit juices | 1.00 | 1.15 (0.74-1.78) | 0.84 (0.53-1.33) | 0.68 (0.41-1.14) | 0.72 (0.45-1.14) | 0.91 (0.55-1.5) | 0.8 (0.51-1.24) | 0.71 (0.43-1.17) | 0.81 (0.5-1.32) | 0.71 (0.43-1.17) | 0.8 (0.66-0.98) | 0.066 |
| Refined grains | 1.00 | 0.99 (0.63-1.53) | 0.79 (0.5-1.26) | 1.04 (0.65-1.66) | 0.68 (0.43-1.09) | 0.72 (0.42-1.22) | 0.59 (0.37-0.93) | 0.82 (0.51-1.3) | 0.78 (0.48-1.28) | 0.78 (0.48-1.25) | 0.82 (0.67-1) | 0.109 |
| Potatoes | 1.00 | 0.93 (0.6-1.46) | 0.75 (0.47-1.19) | 0.7 (0.43-1.15) | 0.63 (0.4-1.01) | 0.95 (0.59-1.54) | 0.8 (0.52-1.24) | 0.53 (0.31-0.88) | 0.75 (0.46-1.21) | 0.68 (0.42-1.1) | 0.82 (0.67-1) | 0.087 |
| Sugar-sweetened beverage | 1.00 | 0.7 (0.45-1.1) | 0.87 (0.57-1.34) | 0.54 (0.32-0.92) | 0.77 (0.5-1.18) | 0.62 (0.37-1.03) | 0.56 (0.35-0.89) | 0.8 (0.51-1.26) | 0.55 (0.33-0.91) | 0.67 (0.42-1.08) | 0.83 (0.68-1.01) | 0.101 |
| Sweets & desserts | 1.00 | 0.79 (0.49-1.26) | 1.19 (0.77-1.83) | 0.85 (0.51-1.39) | 0.76 (0.49-1.2) | 0.61 (0.35-1.05) | 0.65 (0.41-1.03) | 0.95 (0.6-1.49) | 0.69 (0.42-1.14) | 0.88 (0.55-1.42) | 0.86 (0.71-1.05) | 0.294 |
| **Men** | | | | | | | | | | | | |
| Replication | 1.00 | 1.25 (0.84-1.86) | 0.87 (0.55-1.37) | 1 (0.63-1.59) | 1.16 (0.77-1.73) | 0.93 (0.58-1.5) | 0.91 (0.6-1.4) | 1.19 (0.77-1.83) | 1.07 (0.69-1.66) | 1.05 (0.69-1.62) | 1.02 (0.85-1.21) | 0.976 |
| Whole grains | 1.00 | 1.11 (0.74-1.69) | 0.96 (0.63-1.47) | 1.08 (0.68-1.7) | 0.87 (0.57-1.35) | 0.96 (0.6-1.54) | 0.88 (0.58-1.34) | 1.03 (0.63-1.68) | 0.95 (0.62-1.45) | 1.09 (0.71-1.67) | 1 (0.84-1.19) | 0.896 |
| Fruits | 1.00 | 1.27 (0.83-1.94) | 1.24 (0.82-1.88) | 1.27 (0.82-1.97) | 1.02 (0.68-1.55) | 1.12 (0.71-1.78) | 1.12 (0.74-1.68) | 0.92 (0.55-1.53) | 1.14 (0.76-1.71) | 1.35 (0.89-2.05) | 1.08 (0.91-1.29) | 0.492 |
| Vegetables | 1.00 | 1.18 (0.77-1.82) | 1.14 (0.74-1.76) | 1.22 (0.76-1.95) | 1.06 (0.69-1.62) | 1.13 (0.7-1.82) | 1 (0.65-1.53) | 1.26 (0.82-1.96) | 0.94 (0.59-1.49) | 1.11 (0.72-1.74) | 1 (0.84-1.19) | 0.887 |
| Nuts | 1.00 | 0.99 (0.63-1.54) | 1.14 (0.75-1.74) | 1.19 (0.76-1.87) | 1.01 (0.66-1.54) | 1.17 (0.73-1.89) | 0.99 (0.65-1.52) | 1.16 (0.76-1.78) | 0.78 (0.49-1.26) | 1.12 (0.73-1.72) | 1.01 (0.85-1.19) | 0.917 |
| Legumes | 1.00 | 1.16 (0.77-1.74) | 0.83 (0.54-1.27) | 0.97 (0.62-1.49) | 0.83 (0.55-1.24) | 0.97 (0.62-1.53) | 1.04 (0.71-1.52) | 0.94 (0.59-1.49) | 1.05 (0.72-1.54) | 1.06 (0.7-1.59) | 1.05 (0.89-1.25) | 0.641 |
| Vegetables oils | 1.00 | 1.31 (0.85-2.01) | 1.05 (0.67-1.65) | 0.99 (0.61-1.6) | 1.15 (0.75-1.77) | 1.18 (0.72-1.92) | 1.14 (0.74-1.76) | 1.06 (0.67-1.66) | 0.96 (0.6-1.54) | 1.36 (0.88-2.1) | 1.05 (0.88-1.24) | 0.539 |
| Tea & coffee | 1.00 | 1.17 (0.77-1.77) | 1.02 (0.67-1.55) | 0.95 (0.59-1.52) | 0.99 (0.65-1.51) | 1.1 (0.69-1.76) | 1 (0.66-1.51) | 1.25 (0.82-1.92) | 0.91 (0.57-1.43) | 1 (0.65-1.54) | 1.01 (0.86-1.2) | 0.790 |
| Fruit juices | 1.00 | 0.95 (0.63-1.43) | 0.79 (0.51-1.2) | 1.1 (0.72-1.69) | 0.83 (0.55-1.25) | 0.78 (0.49-1.26) | 0.86 (0.57-1.28) | 1.05 (0.7-1.58) | 0.74 (0.47-1.15) | 0.93 (0.62-1.41) | 0.99 (0.84-1.17) | 0.612 |
| Refined grains | 1.00 | 1.18 (0.76-1.83) | 1.24 (0.8-1.91) | 1.05 (0.65-1.7) | 1.2 (0.78-1.83) | 1.01 (0.62-1.64) | 1.06 (0.68-1.64) | 1.15 (0.73-1.8) | 1.22 (0.77-1.92) | 1.2 (0.77-1.86) | 1.04 (0.88-1.23) | 0.624 |
| Potatoes | 1.00 | 1.02 (0.67-1.56) | 1.08 (0.71-1.63) | 1.13 (0.73-1.76) | 0.71 (0.46-1.1) | 0.98 (0.62-1.57) | 0.96 (0.64-1.45) | 1.07 (0.69-1.64) | 0.94 (0.6-1.46) | 1.03 (0.68-1.57) | 0.99 (0.84-1.17) | 0.972 |
| Sugar-sweetened beverage | 1.00 | 0.96 (0.65-1.42) | 0.68 (0.45-1.05) | 0.91 (0.59-1.43) | 0.8 (0.53-1.2) | 0.7 (0.43-1.13) | 0.79 (0.53-1.19) | 1.02 (0.69-1.52) | 0.98 (0.64-1.5) | 0.82 (0.54-1.25) | 0.98 (0.83-1.16) | 0.794 |
| Sweets & desserts | 1.00 | 1.13 (0.74-1.71) | 1.03 (0.67-1.59) | 1.1 (0.7-1.73) | 1.06 (0.7-1.61) | 0.74 (0.45-1.22) | 0.94 (0.62-1.43) | 1.2 (0.79-1.83) | 0.98 (0.63-1.54) | 1.01 (0.66-1.55) | 0.99 (0.84-1.18) | 0.750 |
| **Combined** | | | | | | | | | | | | |
| Replication | 1.00 | 1.04 (0.77-1.39) | 0.89 (0.65-1.23) | 0.8 (0.57-1.13) | 0.94 (0.69-1.26) | 0.76 (0.53-1.08) | 0.81 (0.6-1.1) | 0.94 (0.68-1.29) | 0.89 (0.65-1.23) | 0.84 (0.61-1.16) | 0.93 (0.81-1.06) | 0.220 |
| Whole grains | 1.00 | 0.91 (0.67-1.25) | 0.92 (0.67-1.25) | 1.04 (0.75-1.44) | 0.78 (0.57-1.06) | 0.84 (0.59-1.19) | 0.88 (0.65-1.19) | 0.85 (0.59-1.23) | 0.81 (0.59-1.11) | 0.95 (0.69-1.3) | 0.94 (0.83-1.08) | 0.442 |
| Fruits | 1.00 | 1.14 (0.84-1.56) | 1.11 (0.82-1.5) | 1.06 (0.77-1.47) | 0.84 (0.62-1.15) | 1.03 (0.74-1.43) | 1 (0.75-1.34) | 0.82 (0.57-1.17) | 0.97 (0.72-1.31) | 1.01 (0.74-1.39) | 0.97 (0.85-1.11) | 0.517 |
| Vegetables | 1.00 | 1.05 (0.76-1.44) | 1 (0.73-1.36) | 1.03 (0.73-1.44) | 0.84 (0.61-1.15) | 0.91 (0.64-1.3) | 0.88 (0.65-1.19) | 1.04 (0.75-1.42) | 0.8 (0.57-1.12) | 0.84 (0.6-1.17) | 0.91 (0.8-1.04) | 0.127 |
| Nuts | 1.00 | 0.86 (0.62-1.2) | 0.99 (0.73-1.35) | 1.15 (0.84-1.6) | 0.85 (0.62-1.16) | 0.96 (0.68-1.36) | 0.89 (0.65-1.21) | 0.93 (0.67-1.28) | 0.74 (0.53-1.05) | 0.93 (0.67-1.28) | 0.94 (0.83-1.07) | 0.310 |
| Legumes | 1.00 | 1.1 (0.81-1.49) | 0.84 (0.62-1.15) | 0.84 (0.61-1.17) | 0.83 (0.62-1.11) | 0.91 (0.65-1.26) | 0.91 (0.68-1.22) | 0.83 (0.59-1.17) | 0.94 (0.7-1.26) | 0.86 (0.63-1.17) | 0.95 (0.83-1.08) | 0.372 |
| Vegetables oils | 1.00 | 1.15 (0.84-1.56) | 0.83 (0.59-1.16) | 0.92 (0.65-1.31) | 1.02 (0.74-1.39) | 0.98 (0.7-1.39) | 0.94 (0.69-1.29) | 0.96 (0.69-1.33) | 0.83 (0.59-1.18) | 1.03 (0.74-1.42) | 0.97 (0.85-1.1) | 0.745 |
| Tea & coffee | 1.00 | 0.88 (0.65-1.2) | 0.93 (0.69-1.25) | 0.81 (0.58-1.14) | 0.87 (0.65-1.18) | 0.89 (0.64-1.26) | 0.82 (0.61-1.11) | 0.92 (0.66-1.27) | 0.78 (0.56-1.08) | 0.8 (0.58-1.11) | 0.93 (0.82-1.06) | 0.186 |
| Fruit juices | 1.00 | 1.04 (0.77-1.4) | 0.81 (0.6-1.11) | 0.9 (0.65-1.26) | 0.78 (0.57-1.06) | 0.84 (0.59-1.19) | 0.83 (0.61-1.12) | 0.9 (0.66-1.23) | 0.77 (0.56-1.07) | 0.83 (0.61-1.15) | 0.91 (0.8-1.04) | 0.117 |
| Refined grains | 1.00 | 1.08 (0.79-1.47) | 1.01 (0.73-1.38) | 1.05 (0.75-1.46) | 0.93 (0.68-1.27) | 0.86 (0.6-1.23) | 0.8 (0.58-1.1) | 0.98 (0.71-1.35) | 0.99 (0.71-1.38) | 0.98 (0.71-1.36) | 0.94 (0.83-1.07) | 0.512 |
| Potatoes | 1.00 | 0.98 (0.72-1.33) | 0.92 (0.67-1.24) | 0.92 (0.66-1.27) | 0.67 (0.49-0.92) | 0.97 (0.69-1.35) | 0.89 (0.66-1.19) | 0.8 (0.57-1.11) | 0.85 (0.61-1.17) | 0.86 (0.63-1.18) | 0.92 (0.81-1.04) | 0.265 |
| Sugar-sweetened beverage | 1.00 | 0.84 (0.63-1.13) | 0.77 (0.57-1.05) | 0.74 (0.52-1.04) | 0.78 (0.58-1.05) | 0.66 (0.47-0.94) | 0.68 (0.5-0.93) | 0.92 (0.68-1.24) | 0.77 (0.55-1.06) | 0.75 (0.55-1.03) | 0.91 (0.8-1.04) | 0.211 |
| Sweets & desserts | 1.00 | 0.96 (0.71-1.32) | 1.11 (0.82-1.5) | 0.98 (0.7-1.36) | 0.91 (0.67-1.24) | 0.68 (0.47-0.98) | 0.8 (0.58-1.08) | 1.08 (0.79-1.47) | 0.84 (0.6-1.17) | 0.95 (0.69-1.3) | 0.94 (0.82-1.06) | 0.361 |

Notes: Replication: Per the classification approach advanced by Satija and colleagues (referred to as the scoring approach in the main text). 1) Multivariable-adjusted models were adjusted for age (continuous), multivitamin use, family history of MI (yes vs no), margarine intake (in quintiles), diabetes at baseline, hypercholesterolemia at baseline, hypertension at baseline, updated smoking, updated physical activity, updated alcohol intake, updated calorie intake (in quintiles), updated Aspirin user, updated body mass index category, race, updated Regards region. Models for women also adjusted for hormone use and past oral contraceptive use.

**Supplementary Table 7. Multivariable-adjusted^1^ Hazard ratios and 95% confidence interval for CHD according to deciles of the hPDI (Leave-one-out approach)**

| **Excluded food group** | **Decile 1** | **Decile 2** | **Decile 3** | **Decile 4** | **Decile 5** | **Decile 6** | **Decile 7** | **Decile 8** | **Decile 9** | **Decile 10** | **HR unit=10** | **P trend** |
| --- | --- | --- | --- | --- | --- | --- | --- | --- | --- | --- | --- | --- |
| **Women** | | | | | | | | | | | | |
| Replication | 1.00 | 0.9 (0.57-1.42) | 0.86 (0.55-1.33) | 0.62 (0.38-1.03) | 0.74 (0.46-1.17) | 0.73 (0.46-1.16) | 0.59 (0.36-0.97) | 0.62 (0.37-1.04) | 0.64 (0.39-1.04) | 0.82 (0.51-1.33) | 0.9 (0.76-1.07) | 0.172 |
| Whole grains | 1.00 | 1 (0.64-1.55) | 0.7 (0.43-1.15) | 0.91 (0.57-1.46) | 0.88 (0.55-1.42) | 0.68 (0.42-1.11) | 0.45 (0.26-0.79) | 0.77 (0.47-1.26) | 0.71 (0.43-1.18) | 0.91 (0.56-1.48) | 0.94 (0.8-1.11) | 0.331 |
| Fruits | 1.00 | 0.66 (0.42-1.03) | 0.74 (0.48-1.15) | 0.66 (0.42-1.05) | 0.69 (0.44-1.07) | 0.56 (0.36-0.88) | 0.69 (0.43-1.08) | 0.53 (0.33-0.85) | 0.64 (0.41-1.01) | 0.7 (0.44-1.12) | 0.91 (0.76-1.08) | 0.197 |
| Vegetables | 1.00 | 0.99 (0.64-1.53) | 0.81 (0.51-1.3) | 0.84 (0.52-1.35) | 0.74 (0.46-1.2) | 0.68 (0.42-1.1) | 0.59 (0.36-0.99) | 0.69 (0.42-1.14) | 0.68 (0.42-1.11) | 0.77 (0.46-1.27) | 0.88 (0.74-1.04) | 0.088 |
| Nuts | 1.00 | 0.88 (0.56-1.38) | 0.67 (0.42-1.06) | 0.76 (0.47-1.23) | 0.66 (0.41-1.06) | 0.76 (0.48-1.21) | 0.59 (0.36-0.97) | 0.61 (0.37-1.01) | 0.69 (0.42-1.12) | 0.78 (0.48-1.28) | 0.93 (0.79-1.1) | 0.281 |
| Legumes | 1.00 | 0.95 (0.6-1.51) | 0.93 (0.59-1.44) | 0.89 (0.55-1.42) | 0.68 (0.42-1.11) | 0.69 (0.43-1.11) | 0.64 (0.39-1.05) | 0.74 (0.45-1.23) | 0.69 (0.42-1.14) | 0.82 (0.5-1.33) | 0.9 (0.76-1.07) | 0.152 |
| Vegetables oils | 1.00 | 0.91 (0.58-1.42) | 0.6 (0.38-0.96) | 0.93 (0.59-1.47) | 0.57 (0.35-0.93) | 0.74 (0.47-1.16) | 0.57 (0.34-0.96) | 0.73 (0.45-1.19) | 0.62 (0.38-1.02) | 0.81 (0.51-1.31) | 0.94 (0.79-1.11) | 0.300 |
| Tea & coffee | 1.00 | 0.71 (0.45-1.13) | 0.75 (0.49-1.16) | 0.63 (0.4-0.99) | 0.67 (0.42-1.04) | 0.61 (0.39-0.96) | 0.5 (0.3-0.82) | 0.55 (0.33-0.91) | 0.6 (0.38-0.97) | 0.68 (0.43-1.09) | 0.91 (0.77-1.07) | 0.089 |
| Fruit juices | 1.00 | 0.88 (0.57-1.36) | 0.65 (0.41-1.04) | 0.76 (0.49-1.19) | 0.56 (0.35-0.9) | 0.68 (0.44-1.06) | 0.75 (0.48-1.16) | 0.58 (0.36-0.93) | 0.47 (0.28-0.77) | 0.85 (0.55-1.32) | 0.91 (0.77-1.08) | 0.187 |
| Refined grains | 1.00 | 0.82 (0.52-1.29) | 0.78 (0.49-1.25) | 0.97 (0.63-1.49) | 0.62 (0.38-0.99) | 0.67 (0.4-1.11) | 0.64 (0.4-1.03) | 0.83 (0.53-1.3) | 0.51 (0.32-0.82) | 0.86 (0.54-1.35) | 0.9 (0.76-1.07) | 0.153 |
| Potatoes | 1.00 | 0.77 (0.49-1.19) | 0.68 (0.43-1.09) | 0.68 (0.43-1.09) | 0.66 (0.42-1.02) | 0.81 (0.5-1.32) | 0.69 (0.44-1.09) | 0.54 (0.34-0.85) | 0.56 (0.35-0.91) | 0.81 (0.51-1.29) | 0.9 (0.76-1.06) | 0.210 |
| Sugar-sweetened beverage | 1.00 | 0.91 (0.59-1.4) | 0.85 (0.54-1.32) | 0.51 (0.31-0.84) | 0.79 (0.51-1.22) | 0.72 (0.44-1.17) | 0.61 (0.39-0.98) | 0.63 (0.4-0.98) | 0.53 (0.32-0.88) | 0.8 (0.5-1.26) | 0.89 (0.75-1.06) | 0.093 |
| Sweets & desserts | 1.00 | 1.03 (0.67-1.58) | 0.76 (0.47-1.22) | 0.89 (0.57-1.37) | 0.72 (0.45-1.16) | 0.53 (0.3-0.93) | 0.72 (0.45-1.14) | 0.7 (0.43-1.12) | 0.55 (0.34-0.89) | 0.84 (0.53-1.34) | 0.87 (0.73-1.03) | 0.061 |
| **Men** | | | | | | | | | | | | |
| Replication | 1.00 | 1.03 (0.68-1.57) | 0.83 (0.55-1.26) | 0.93 (0.61-1.41) | 0.67 (0.42-1.06) | 0.84 (0.56-1.28) | 0.8 (0.52-1.24) | 0.9 (0.57-1.4) | 0.61 (0.38-0.97) | 0.8 (0.51-1.26) | 0.89 (0.76-1.03) | 0.089 |
| Whole grains | 1.00 | 0.86 (0.57-1.3) | 0.88 (0.57-1.37) | 1.07 (0.7-1.63) | 0.67 (0.42-1.04) | 0.76 (0.49-1.18) | 0.85 (0.55-1.33) | 0.61 (0.38-0.99) | 0.73 (0.46-1.15) | 0.79 (0.49-1.26) | 0.87 (0.75-1.01) | 0.132 |
| Fruits | 1.00 | 0.68 (0.44-1.04) | 0.82 (0.54-1.23) | 1.02 (0.68-1.52) | 0.82 (0.55-1.22) | 0.64 (0.41-0.98) | 0.93 (0.62-1.39) | 0.78 (0.51-1.21) | 0.65 (0.41-1.01) | 0.85 (0.55-1.31) | 0.92 (0.79-1.07) | 0.459 |
| Vegetables | 1.00 | 0.97 (0.64-1.47) | 1.02 (0.67-1.56) | 1.09 (0.71-1.67) | 0.72 (0.46-1.12) | 0.8 (0.51-1.25) | 0.84 (0.54-1.32) | 0.95 (0.61-1.49) | 0.64 (0.4-1.05) | 0.83 (0.52-1.33) | 0.87 (0.74-1.01) | 0.112 |
| Nuts | 1.00 | 1 (0.66-1.51) | 0.71 (0.46-1.08) | 1.01 (0.65-1.55) | 0.62 (0.39-0.98) | 0.84 (0.55-1.3) | 0.78 (0.5-1.22) | 0.79 (0.5-1.26) | 0.62 (0.38-0.99) | 0.74 (0.46-1.19) | 0.87 (0.75-1.02) | 0.112 |
| Legumes | 1.00 | 0.82 (0.53-1.27) | 0.87 (0.58-1.31) | 0.85 (0.54-1.32) | 0.72 (0.47-1.11) | 0.88 (0.58-1.35) | 0.82 (0.52-1.28) | 0.93 (0.6-1.45) | 0.58 (0.36-0.93) | 0.77 (0.49-1.22) | 0.91 (0.78-1.05) | 0.177 |
| Vegetables oils | 1.00 | 0.8 (0.5-1.26) | 1.15 (0.77-1.74) | 1.04 (0.67-1.61) | 0.73 (0.46-1.16) | 0.87 (0.56-1.35) | 0.86 (0.54-1.36) | 0.86 (0.54-1.37) | 0.74 (0.46-1.19) | 0.84 (0.52-1.36) | 0.91 (0.78-1.05) | 0.191 |
| Tea & coffee | 1.00 | 0.98 (0.64-1.5) | 0.96 (0.64-1.45) | 0.93 (0.61-1.42) | 0.86 (0.56-1.32) | 0.83 (0.54-1.27) | 0.85 (0.55-1.32) | 0.82 (0.52-1.28) | 0.77 (0.48-1.22) | 0.81 (0.51-1.28) | 0.89 (0.76-1.03) | 0.163 |
| Fruit juices | 1.00 | 1.25 (0.83-1.9) | 1.17 (0.77-1.76) | 1.01 (0.66-1.56) | 0.84 (0.54-1.3) | 0.99 (0.65-1.52) | 1 (0.65-1.55) | 0.89 (0.56-1.4) | 1.09 (0.71-1.68) | 0.86 (0.54-1.38) | 0.91 (0.78-1.05) | 0.284 |
| Refined grains | 1.00 | 1 (0.66-1.5) | 0.84 (0.54-1.29) | 0.9 (0.59-1.38) | 0.76 (0.49-1.17) | 0.86 (0.54-1.35) | 0.78 (0.51-1.19) | 0.95 (0.62-1.45) | 0.71 (0.45-1.1) | 0.77 (0.49-1.2) | 0.87 (0.75-1.01) | 0.137 |
| Potatoes | 1.00 | 0.98 (0.65-1.47) | 0.87 (0.56-1.35) | 0.89 (0.58-1.37) | 0.99 (0.66-1.48) | 0.82 (0.51-1.3) | 0.92 (0.61-1.39) | 0.76 (0.49-1.15) | 0.88 (0.57-1.35) | 0.85 (0.54-1.34) | 0.9 (0.78-1.05) | 0.321 |
| Sugar-sweetened beverage | 1.00 | 1.13 (0.74-1.73) | 1.2 (0.78-1.85) | 0.94 (0.6-1.47) | 1.07 (0.7-1.63) | 0.87 (0.53-1.43) | 0.92 (0.59-1.42) | 0.99 (0.65-1.5) | 0.86 (0.54-1.38) | 0.97 (0.62-1.54) | 0.91 (0.78-1.06) | 0.331 |
| Sweets & desserts | 1.00 | 1.29 (0.86-1.96) | 0.84 (0.53-1.35) | 1.26 (0.83-1.92) | 0.86 (0.55-1.35) | 1.13 (0.71-1.79) | 1.01 (0.66-1.55) | 0.99 (0.64-1.55) | 0.96 (0.63-1.49) | 0.89 (0.56-1.41) | 0.9 (0.78-1.04) | 0.229 |
| **Combined** | | | | | | | | | | | | |
| Replication | 1.00 | 0.97 (0.71-1.32) | 0.84 (0.62-1.14) | 0.79 (0.57-1.09) | 0.7 (0.51-0.97) | 0.79 (0.58-1.08) | 0.7 (0.51-0.97) | 0.77 (0.55-1.08) | 0.62 (0.44-0.87) | 0.81 (0.58-1.13) | 0.89 (0.8-1) | 0.029 |
| Whole grains | 1.00 | 0.93 (0.69-1.25) | 0.8 (0.57-1.11) | 0.99 (0.73-1.36) | 0.76 (0.55-1.06) | 0.72 (0.52-1) | 0.67 (0.47-0.94) | 0.68 (0.48-0.96) | 0.72 (0.51-1.01) | 0.85 (0.6-1.19) | 0.9 (0.8-1.01) | 0.078 |
| Fruits | 1.00 | 0.67 (0.49-0.91) | 0.78 (0.58-1.05) | 0.85 (0.63-1.14) | 0.76 (0.56-1.01) | 0.6 (0.44-0.82) | 0.81 (0.6-1.1) | 0.65 (0.47-0.9) | 0.64 (0.47-0.88) | 0.78 (0.56-1.07) | 0.92 (0.82-1.03) | 0.156 |
| Vegetables | 1.00 | 0.98 (0.72-1.32) | 0.92 (0.67-1.27) | 0.97 (0.7-1.33) | 0.73 (0.53-1.01) | 0.74 (0.54-1.03) | 0.72 (0.52-1.01) | 0.83 (0.59-1.15) | 0.66 (0.47-0.93) | 0.8 (0.57-1.13) | 0.87 (0.78-0.98) | 0.020 |
| Nuts | 1.00 | 0.94 (0.69-1.28) | 0.69 (0.51-0.94) | 0.89 (0.64-1.23) | 0.64 (0.46-0.89) | 0.8 (0.59-1.1) | 0.69 (0.49-0.96) | 0.7 (0.5-0.99) | 0.65 (0.46-0.91) | 0.76 (0.54-1.07) | 0.9 (0.8-1.01) | 0.057 |
| Legumes | 1.00 | 0.88 (0.64-1.21) | 0.9 (0.67-1.21) | 0.87 (0.63-1.2) | 0.7 (0.51-0.97) | 0.79 (0.58-1.09) | 0.73 (0.53-1.02) | 0.84 (0.61-1.18) | 0.63 (0.44-0.89) | 0.79 (0.57-1.11) | 0.91 (0.81-1.01) | 0.050 |
| Vegetables oils | 1.00 | 0.85 (0.62-1.17) | 0.87 (0.64-1.19) | 0.99 (0.72-1.35) | 0.65 (0.46-0.91) | 0.8 (0.58-1.1) | 0.72 (0.51-1.02) | 0.8 (0.57-1.11) | 0.68 (0.48-0.96) | 0.83 (0.59-1.16) | 0.92 (0.82-1.03) | 0.096 |
| Tea & coffee | 1.00 | 0.84 (0.62-1.15) | 0.86 (0.64-1.15) | 0.78 (0.57-1.06) | 0.76 (0.56-1.04) | 0.72 (0.53-0.98) | 0.67 (0.48-0.94) | 0.68 (0.49-0.96) | 0.68 (0.49-0.95) | 0.75 (0.54-1.03) | 0.89 (0.8-1) | 0.030 |
| Fruit juices | 1.00 | 1.06 (0.79-1.43) | 0.9 (0.66-1.23) | 0.88 (0.65-1.2) | 0.7 (0.51-0.96) | 0.83 (0.61-1.12) | 0.86 (0.63-1.18) | 0.72 (0.52-1) | 0.76 (0.55-1.06) | 0.86 (0.62-1.18) | 0.91 (0.81-1.01) | 0.094 |
| Refined grains | 1.00 | 0.91 (0.67-1.24) | 0.81 (0.59-1.11) | 0.93 (0.69-1.26) | 0.69 (0.5-0.95) | 0.77 (0.55-1.07) | 0.72 (0.52-0.98) | 0.89 (0.66-1.21) | 0.61 (0.44-0.84) | 0.81 (0.59-1.12) | 0.88 (0.79-0.99) | 0.039 |
| Potatoes | 1.00 | 0.87 (0.64-1.18) | 0.78 (0.57-1.07) | 0.79 (0.57-1.08) | 0.82 (0.61-1.1) | 0.82 (0.58-1.14) | 0.81 (0.59-1.09) | 0.65 (0.48-0.88) | 0.72 (0.52-0.99) | 0.83 (0.6-1.15) | 0.9 (0.8-1.01) | 0.116 |
| Sugar-sweetened beverage | 1.00 | 1.02 (0.75-1.37) | 1.01 (0.74-1.38) | 0.71 (0.51-1) | 0.92 (0.68-1.25) | 0.79 (0.56-1.12) | 0.76 (0.55-1.04) | 0.8 (0.59-1.08) | 0.69 (0.49-0.97) | 0.88 (0.64-1.22) | 0.9 (0.81-1.01) | 0.066 |
| Sweets & desserts | 1.00 | 1.16 (0.86-1.56) | 0.8 (0.57-1.12) | 1.07 (0.79-1.44) | 0.79 (0.57-1.1) | 0.83 (0.58-1.19) | 0.87 (0.63-1.18) | 0.84 (0.61-1.17) | 0.75 (0.54-1.03) | 0.87 (0.62-1.2) | 0.89 (0.79-0.99) | 0.033 |

Notes: Replication: Per the classification approach advanced by Satija and colleagues (referred to as the scoring approach in the main text). 1) Multivariable-adjusted models were adjusted for age (continuous), multivitamin use, family history of MI (yes vs no), margarine intake (in quintiles), diabetes at baseline, hypercholesterolemia at baseline, hypertension at baseline, updated smoking, updated physical activity, updated alcohol intake, updated calorie intake (in quintiles), updated Aspirin user, updated body mass index category, race, updated Regards region. Models for women also adjusted for hormone use and past oral contraceptive use.

**Supplementary Table 8. Multivariable-adjusted^1^ Hazard ratios and 95% confidence interval for CHD according to deciles of the uPDI (****Leave-one-out approach)**

| **Excluded food group** | **Decile 1** | **Decile 2** | **Decile 3** | **Decile 4** | **Decile 5** | **Decile 6** | **Decile 7** | **Decile 8** | **Decile 9** | **Decile 10** | **HR unit=10** | **P trend** |
| --- | --- | --- | --- | --- | --- | --- | --- | --- | --- | --- | --- | --- |
| **Women** | | | | | | | | | | | | |
| Replication | 1.00 | 1.24 (0.74-2.07) | 1.21 (0.73-2.02) | 1.14 (0.67-1.94) | 1.15 (0.68-1.95) | 0.94 (0.54-1.62) | 1.31 (0.78-2.22) | 1.41 (0.84-2.37) | 1.27 (0.74-2.16) | 1.41 (0.83-2.38) | 1.1 (0.94-1.29) | 0.198 |
| Whole grains | 1.00 | 1.18 (0.73-1.91) | 0.81 (0.49-1.33) | 1.22 (0.76-1.95) | 0.71 (0.42-1.21) | 0.84 (0.51-1.38) | 0.94 (0.57-1.55) | 1.15 (0.7-1.88) | 1.15 (0.71-1.85) | 0.98 (0.59-1.6) | 1.06 (0.91-1.25) | 0.822 |
| Fruits | 1.00 | 1.33 (0.81-2.18) | 0.85 (0.51-1.43) | 1.28 (0.79-2.09) | 0.89 (0.52-1.51) | 0.92 (0.55-1.56) | 1.09 (0.65-1.81) | 1.2 (0.72-2) | 1.35 (0.81-2.23) | 1.27 (0.77-2.12) | 1.1 (0.93-1.29) | 0.278 |
| Vegetables | 1.00 | 1.13 (0.7-1.84) | 1.09 (0.66-1.81) | 1.04 (0.63-1.72) | 1.01 (0.62-1.65) | 1.11 (0.68-1.79) | 1.07 (0.65-1.76) | 1.31 (0.8-2.16) | 1.23 (0.75-2) | 1.35 (0.82-2.23) | 1.13 (0.96-1.34) | 0.160 |
| Nuts | 1.00 | 1.34 (0.82-2.19) | 1.1 (0.67-1.79) | 1.07 (0.64-1.79) | 1.05 (0.63-1.74) | 0.79 (0.46-1.35) | 1.25 (0.76-2.05) | 1.27 (0.76-2.12) | 1.24 (0.75-2.05) | 1.18 (0.71-1.96) | 1.07 (0.92-1.25) | 0.513 |
| Legumes | 1.00 | 1.04 (0.62-1.74) | 1.16 (0.71-1.87) | 0.95 (0.56-1.6) | 1.07 (0.65-1.77) | 0.92 (0.55-1.53) | 1.2 (0.73-1.98) | 1.14 (0.68-1.92) | 1.13 (0.68-1.88) | 1.32 (0.8-2.19) | 1.11 (0.94-1.31) | 0.254 |
| Vegetables oils | 1.00 | 1.02 (0.63-1.66) | 1.1 (0.69-1.75) | 1.02 (0.62-1.67) | 0.74 (0.44-1.24) | 0.88 (0.53-1.47) | 1.18 (0.73-1.9) | 1.24 (0.76-2.01) | 0.92 (0.56-1.52) | 1.24 (0.75-2.03) | 1.07 (0.91-1.25) | 0.479 |
| Tea & coffee | 1.00 | 1.04 (0.64-1.69) | 0.89 (0.55-1.46) | 1.13 (0.69-1.83) | 0.78 (0.47-1.3) | 1.04 (0.64-1.7) | 1.04 (0.64-1.71) | 1.18 (0.72-1.93) | 1.15 (0.71-1.86) | 1.15 (0.7-1.89) | 1.1 (0.94-1.28) | 0.299 |
| Fruit juices | 1.00 | 0.9 (0.56-1.46) | 0.81 (0.51-1.29) | 0.75 (0.46-1.23) | 0.91 (0.57-1.46) | 0.99 (0.62-1.58) | 1.08 (0.68-1.71) | 1.03 (0.64-1.65) | 0.84 (0.51-1.38) | 1.11 (0.69-1.77) | 1.09 (0.93-1.27) | 0.365 |
| Refined grains | 1.00 | 1.01 (0.61-1.67) | 1.03 (0.6-1.78) | 1.39 (0.86-2.24) | 0.85 (0.5-1.47) | 1.13 (0.68-1.89) | 1.25 (0.74-2.09) | 1.14 (0.69-1.9) | 1.24 (0.73-2.1) | 1.25 (0.74-2.12) | 1.1 (0.94-1.28) | 0.331 |
| Potatoes | 1.00 | 0.62 (0.37-1.02) | 0.92 (0.56-1.5) | 0.91 (0.58-1.42) | 0.71 (0.43-1.18) | 0.86 (0.53-1.39) | 0.92 (0.55-1.52) | 1.01 (0.63-1.61) | 0.92 (0.56-1.51) | 1.02 (0.62-1.67) | 1.1 (0.94-1.28) | 0.347 |
| Sugar-sweetened beverage | 1.00 | 0.81 (0.49-1.36) | 0.9 (0.55-1.46) | 0.91 (0.55-1.51) | 1.19 (0.75-1.89) | 0.99 (0.61-1.61) | 0.73 (0.43-1.24) | 1.09 (0.66-1.79) | 1.16 (0.72-1.86) | 1.07 (0.64-1.77) | 1.11 (0.95-1.31) | 0.326 |
| Sweets & desserts | 1.00 | 1.26 (0.76-2.08) | 0.81 (0.47-1.4) | 1.3 (0.78-2.17) | 1.02 (0.6-1.74) | 1.4 (0.84-2.34) | 1.19 (0.69-2.05) | 1.2 (0.71-2.01) | 1.39 (0.81-2.38) | 1.36 (0.79-2.33) | 1.14 (0.97-1.33) | 0.136 |
| **Men** | | | | | | | | | | | | |
| Replication | 1.00 | 0.85 (0.54-1.33) | 0.93 (0.6-1.44) | 0.88 (0.55-1.4) | 1.07 (0.69-1.65) | 0.87 (0.55-1.37) | 1.02 (0.65-1.6) | 1.16 (0.75-1.8) | 1.12 (0.71-1.78) | 1.05 (0.66-1.68) | 1.12 (0.97-1.3) | 0.286 |
| Whole grains | 1.00 | 0.91 (0.57-1.44) | 0.97 (0.63-1.49) | 0.91 (0.58-1.43) | 1.15 (0.75-1.76) | 0.9 (0.58-1.4) | 0.96 (0.6-1.52) | 1.08 (0.69-1.7) | 1.41 (0.92-2.18) | 1.18 (0.75-1.87) | 1.14 (0.99-1.32) | 0.093 |
| Fruits | 1.00 | 0.81 (0.51-1.3) | 0.82 (0.53-1.27) | 1.06 (0.68-1.64) | 1.03 (0.67-1.6) | 1.04 (0.68-1.59) | 0.99 (0.63-1.54) | 1.18 (0.76-1.83) | 0.89 (0.56-1.42) | 1.09 (0.69-1.72) | 1.08 (0.94-1.25) | 0.355 |
| Vegetables | 1.00 | 0.94 (0.6-1.45) | 1.1 (0.7-1.72) | 0.83 (0.53-1.31) | 1.05 (0.69-1.6) | 1.04 (0.67-1.6) | 1.08 (0.7-1.67) | 1.48 (0.97-2.26) | 1.1 (0.7-1.72) | 1.19 (0.75-1.87) | 1.15 (0.99-1.33) | 0.154 |
| Nuts | 1.00 | 1.04 (0.65-1.65) | 0.97 (0.63-1.5) | 0.99 (0.62-1.56) | 1.04 (0.67-1.62) | 1.18 (0.77-1.8) | 1.04 (0.67-1.63) | 1.33 (0.85-2.08) | 1.22 (0.79-1.89) | 1.3 (0.83-2.03) | 1.13 (0.98-1.3) | 0.092 |
| Legumes | 1.00 | 0.82 (0.51-1.31) | 0.81 (0.52-1.25) | 1 (0.64-1.55) | 0.96 (0.62-1.48) | 0.88 (0.57-1.35) | 0.81 (0.51-1.28) | 1.63 (1.08-2.46) | 0.79 (0.5-1.26) | 1.06 (0.67-1.67) | 1.1 (0.95-1.28) | 0.306 |
| Vegetables oils | 1.00 | 0.92 (0.58-1.44) | 0.96 (0.63-1.48) | 0.96 (0.62-1.5) | 1.21 (0.79-1.86) | 0.84 (0.54-1.31) | 1.05 (0.67-1.63) | 1.11 (0.71-1.74) | 1.19 (0.78-1.81) | 1.13 (0.7-1.83) | 1.1 (0.96-1.27) | 0.243 |
| Tea & coffee | 1.00 | 0.85 (0.54-1.34) | 0.95 (0.62-1.45) | 0.84 (0.53-1.35) | 0.86 (0.55-1.35) | 1.08 (0.7-1.66) | 0.97 (0.63-1.51) | 0.95 (0.6-1.51) | 1.25 (0.82-1.91) | 1.13 (0.71-1.79) | 1.12 (0.98-1.29) | 0.142 |
| Fruit juices | 1.00 | 0.8 (0.51-1.24) | 0.95 (0.62-1.45) | 1.07 (0.7-1.62) | 0.83 (0.53-1.31) | 1.15 (0.75-1.74) | 0.81 (0.51-1.28) | 0.95 (0.62-1.46) | 1.02 (0.65-1.6) | 1.2 (0.77-1.89) | 1.1 (0.96-1.27) | 0.318 |
| Refined grains | 1.00 | 0.95 (0.6-1.5) | 1.35 (0.86-2.12) | 0.93 (0.59-1.47) | 1.12 (0.71-1.77) | 1.06 (0.66-1.7) | 1.09 (0.68-1.75) | 1.47 (0.95-2.29) | 1.27 (0.79-2.04) | 1.29 (0.79-2.09) | 1.14 (0.99-1.31) | 0.099 |
| Potatoes | 1.00 | 0.94 (0.6-1.48) | 1.26 (0.79-2.01) | 1.06 (0.7-1.62) | 0.95 (0.59-1.52) | 1.14 (0.73-1.78) | 1.2 (0.76-1.9) | 1.19 (0.76-1.86) | 1.32 (0.83-2.09) | 1.12 (0.7-1.82) | 1.1 (0.96-1.26) | 0.316 |
| Sugar-sweetened beverage | 1.00 | 1.03 (0.66-1.61) | 0.95 (0.61-1.48) | 1.09 (0.7-1.71) | 1.05 (0.67-1.64) | 1.08 (0.69-1.69) | 1.05 (0.67-1.65) | 1.24 (0.79-1.95) | 1.22 (0.79-1.89) | 1.18 (0.73-1.91) | 1.1 (0.95-1.27) | 0.211 |
| Sweets & desserts | 1.00 | 1.1 (0.69-1.74) | 1.16 (0.73-1.85) | 0.99 (0.61-1.61) | 1.03 (0.64-1.67) | 1.31 (0.82-2.1) | 1.12 (0.68-1.83) | 1.4 (0.88-2.24) | 1.14 (0.68-1.91) | 1.34 (0.81-2.22) | 1.11 (0.96-1.27) | 0.175 |
| **Combined** | | | | | | | | | | | | |
| Replication | 1.00 | 1 (0.71-1.4) | 1.04 (0.75-1.45) | 0.98 (0.69-1.4) | 1.1 (0.79-1.54) | 0.9 (0.63-1.27) | 1.14 (0.81-1.6) | 1.26 (0.9-1.76) | 1.18 (0.84-1.68) | 1.19 (0.84-1.69) | 1.11 (1-1.24) | 0.098 |
| Whole grains | 1.00 | 1.03 (0.74-1.44) | 0.89 (0.65-1.24) | 1.05 (0.76-1.45) | 0.95 (0.68-1.33) | 0.87 (0.63-1.21) | 0.95 (0.68-1.33) | 1.11 (0.8-1.55) | 1.29 (0.93-1.77) | 1.08 (0.77-1.52) | 1.11 (0.99-1.23) | 0.163 |
| Fruits | 1.00 | 1.02 (0.73-1.44) | 0.83 (0.6-1.16) | 1.16 (0.83-1.6) | 0.97 (0.7-1.36) | 0.99 (0.71-1.38) | 1.03 (0.74-1.44) | 1.19 (0.85-1.66) | 1.08 (0.77-1.51) | 1.17 (0.83-1.64) | 1.09 (0.98-1.21) | 0.159 |
| Vegetables | 1.00 | 1.02 (0.74-1.41) | 1.09 (0.78-1.53) | 0.92 (0.66-1.29) | 1.03 (0.75-1.42) | 1.07 (0.77-1.47) | 1.08 (0.78-1.5) | 1.41 (1.02-1.94) | 1.15 (0.83-1.61) | 1.26 (0.9-1.76) | 1.14 (1.02-1.28) | 0.046 |
| Nuts | 1.00 | 1.17 (0.84-1.64) | 1.03 (0.74-1.42) | 1.02 (0.73-1.44) | 1.05 (0.75-1.46) | 1.01 (0.72-1.41) | 1.13 (0.81-1.57) | 1.3 (0.93-1.83) | 1.23 (0.89-1.71) | 1.25 (0.89-1.74) | 1.1 (0.99-1.22) | 0.091 |
| Legumes | 1.00 | 0.91 (0.64-1.29) | 0.95 (0.69-1.31) | 0.98 (0.7-1.37) | 1 (0.72-1.39) | 0.9 (0.64-1.25) | 0.97 (0.69-1.36) | 1.42 (1.03-1.96) | 0.93 (0.66-1.32) | 1.17 (0.84-1.64) | 1.11 (0.99-1.24) | 0.128 |
| Vegetables oils | 1.00 | 0.96 (0.69-1.34) | 1.03 (0.75-1.4) | 0.99 (0.71-1.37) | 1 (0.72-1.38) | 0.86 (0.61-1.2) | 1.11 (0.8-1.53) | 1.17 (0.84-1.62) | 1.07 (0.77-1.47) | 1.18 (0.84-1.67) | 1.09 (0.98-1.21) | 0.180 |
| Tea & coffee | 1.00 | 0.93 (0.67-1.3) | 0.93 (0.67-1.28) | 0.97 (0.69-1.36) | 0.83 (0.59-1.16) | 1.06 (0.77-1.47) | 1 (0.72-1.39) | 1.05 (0.75-1.47) | 1.21 (0.88-1.66) | 1.14 (0.81-1.6) | 1.11 (1-1.23) | 0.074 |
| Fruit juices | 1.00 | 0.85 (0.61-1.17) | 0.88 (0.65-1.21) | 0.92 (0.67-1.26) | 0.87 (0.63-1.2) | 1.07 (0.79-1.47) | 0.94 (0.68-1.3) | 0.98 (0.72-1.35) | 0.94 (0.67-1.31) | 1.16 (0.83-1.6) | 1.1 (0.99-1.22) | 0.177 |
| Refined grains | 1.00 | 0.98 (0.69-1.37) | 1.21 (0.86-1.71) | 1.13 (0.81-1.57) | 1 (0.71-1.42) | 1.09 (0.77-1.55) | 1.16 (0.82-1.65) | 1.32 (0.95-1.84) | 1.26 (0.88-1.79) | 1.27 (0.89-1.81) | 1.12 (1.01-1.24) | 0.061 |
| Potatoes | 1.00 | 0.78 (0.56-1.09) | 1.08 (0.77-1.52) | 0.99 (0.73-1.34) | 0.83 (0.59-1.17) | 1 (0.72-1.38) | 1.06 (0.76-1.49) | 1.1 (0.79-1.52) | 1.11 (0.8-1.56) | 1.07 (0.76-1.51) | 1.1 (0.99-1.22) | 0.169 |
| Sugar-sweetened beverage | 1.00 | 0.93 (0.66-1.3) | 0.93 (0.67-1.28) | 1.01 (0.72-1.41) | 1.11 (0.81-1.53) | 1.04 (0.75-1.44) | 0.9 (0.64-1.27) | 1.17 (0.84-1.63) | 1.19 (0.86-1.65) | 1.13 (0.79-1.6) | 1.11 (0.99-1.23) | 0.113 |
| Sweets & desserts | 1.00 | 1.17 (0.83-1.64) | 1 (0.7-1.42) | 1.13 (0.79-1.61) | 1.03 (0.72-1.47) | 1.35 (0.96-1.91) | 1.15 (0.8-1.66) | 1.31 (0.92-1.85) | 1.25 (0.86-1.82) | 1.35 (0.93-1.95) | 1.12 (1.01-1.24) | 0.045 |

Notes: Replication: Per the classification approach advanced by Satija and colleagues (referred to as the scoring approach in the main text). 1) Multivariable-adjusted models were adjusted for age (continuous), multivitamin use, family history of MI (yes vs no), margarine intake (in quintiles), diabetes at baseline, hypercholesterolemia at baseline, hypertension at baseline, updated smoking, updated physical activity, updated alcohol intake, updated calorie intake (in quintiles), updated Aspirin user, updated body mass index category, race, updated Regards region. Models for women also adjusted for hormone use and past oral contraceptive use.

**Supplementary Figure 1.** Sample size flow diagram

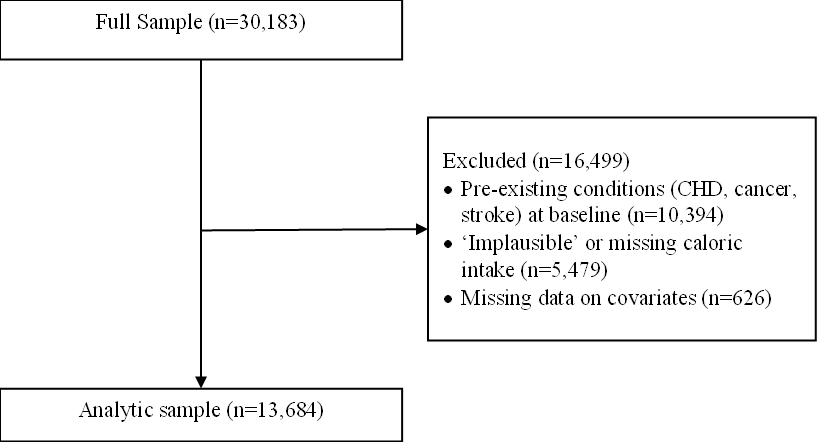

**Supplementary Figure 2.** Means of diet indices at baseline (Recategorization approach)

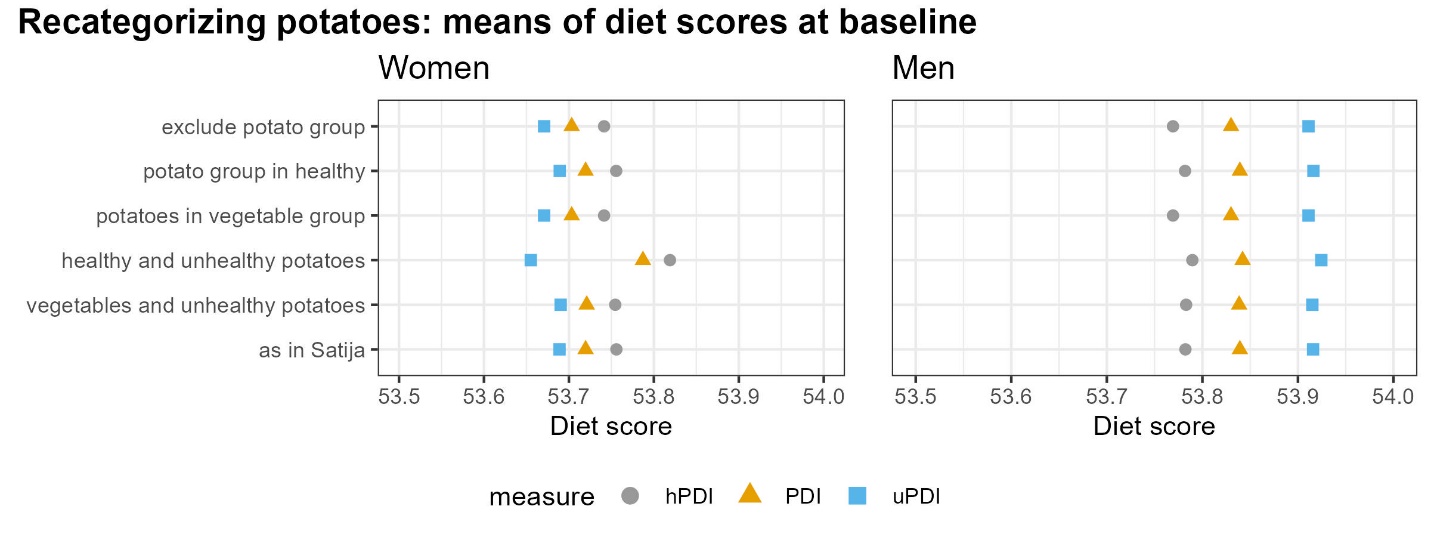

**Supplementary Figure 3.** Means of diet indices at baseline (Leave-one-out approach)

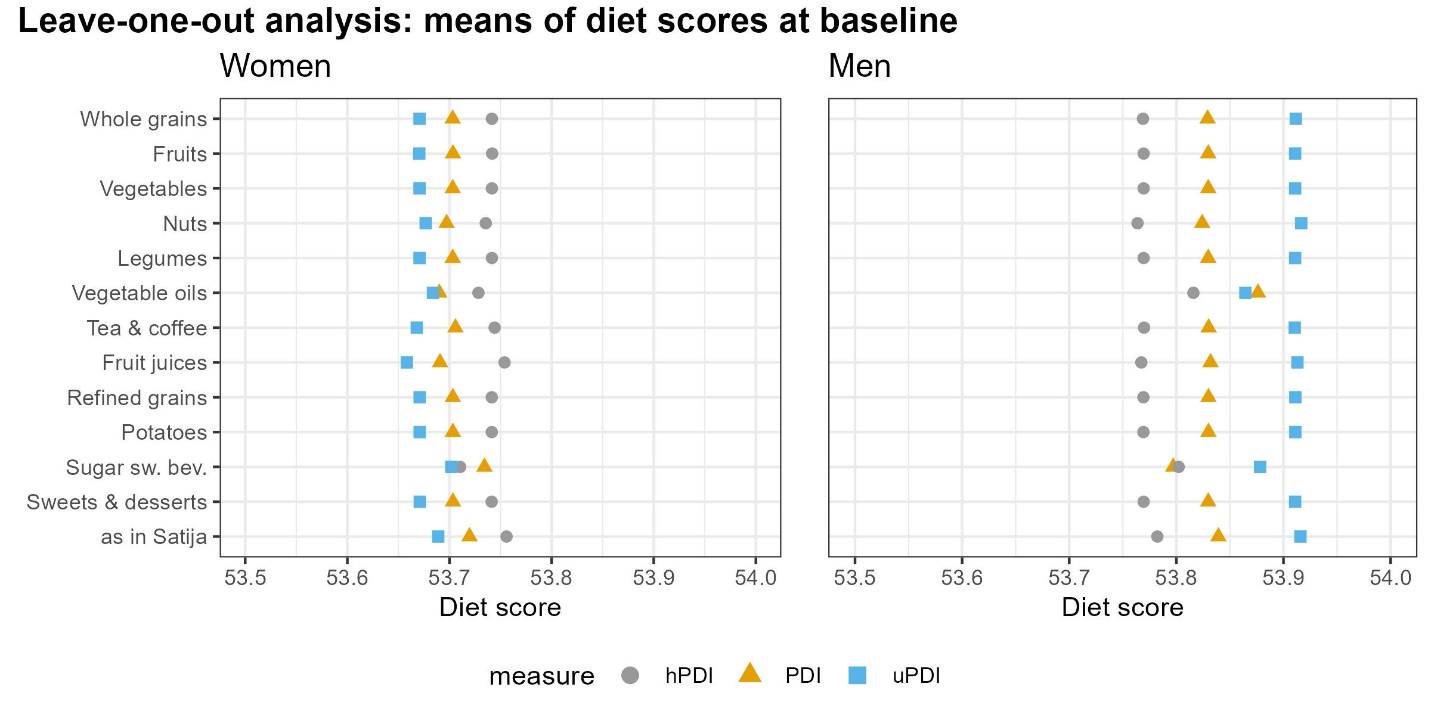

**Supplementary Figure 4.** Agnostic permutated models where each of the 12 plant-based food groups are either positively (i.e., more consumption is better; depicted in orange) or reversely (less consumption is better, depicted in blue) scored, using all combinations (violin plots showing the distributions of hazard ratios of the highest decile compared to the lowest decile of hPDI).

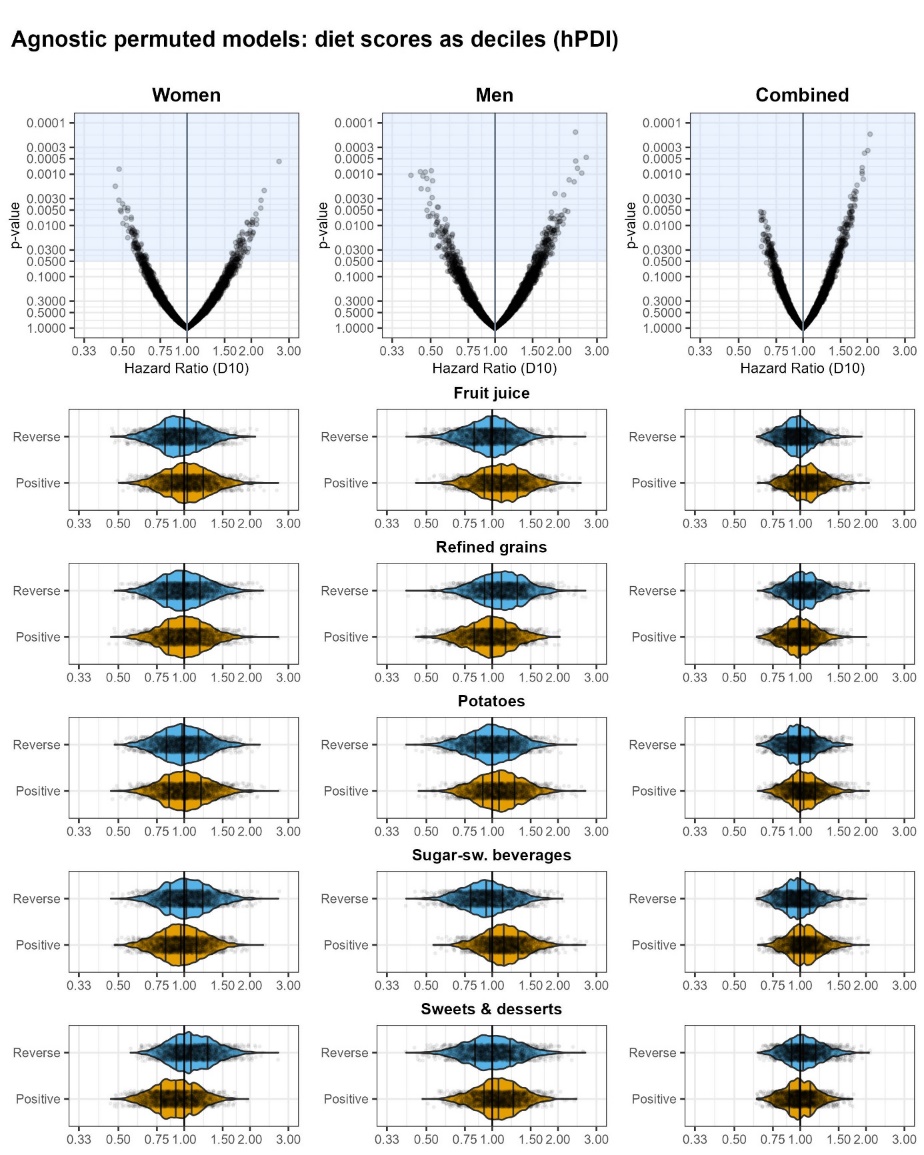

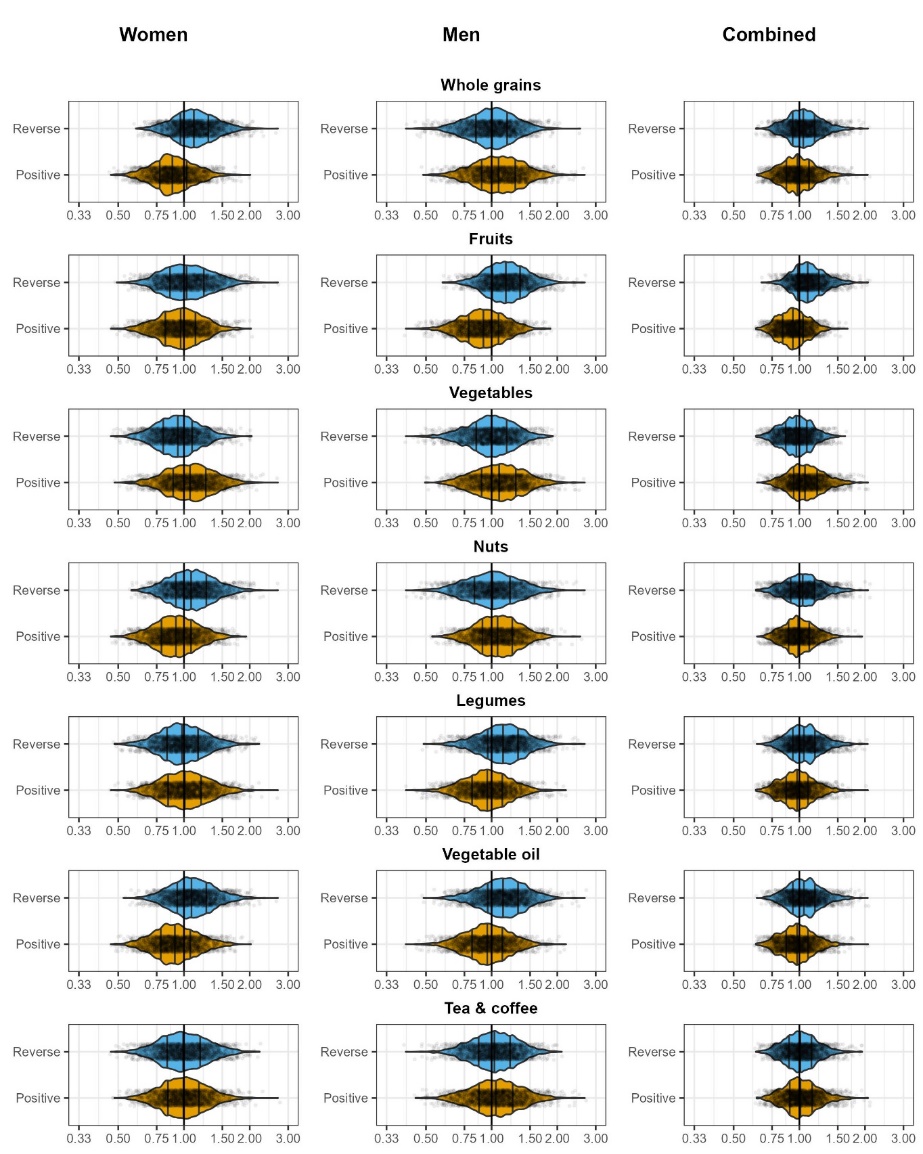

**Supplementary Figure 5.** Agnostic permuted models where each of the 12 plant-based food groups are either positively (i.e., more consumption is better; depicted in orange) or reversely (less consumption is better, depicted in blue) scored, using all combinations (empirical cumulative density function (ECDF) of hazard ratios for continuous hPDI scores).

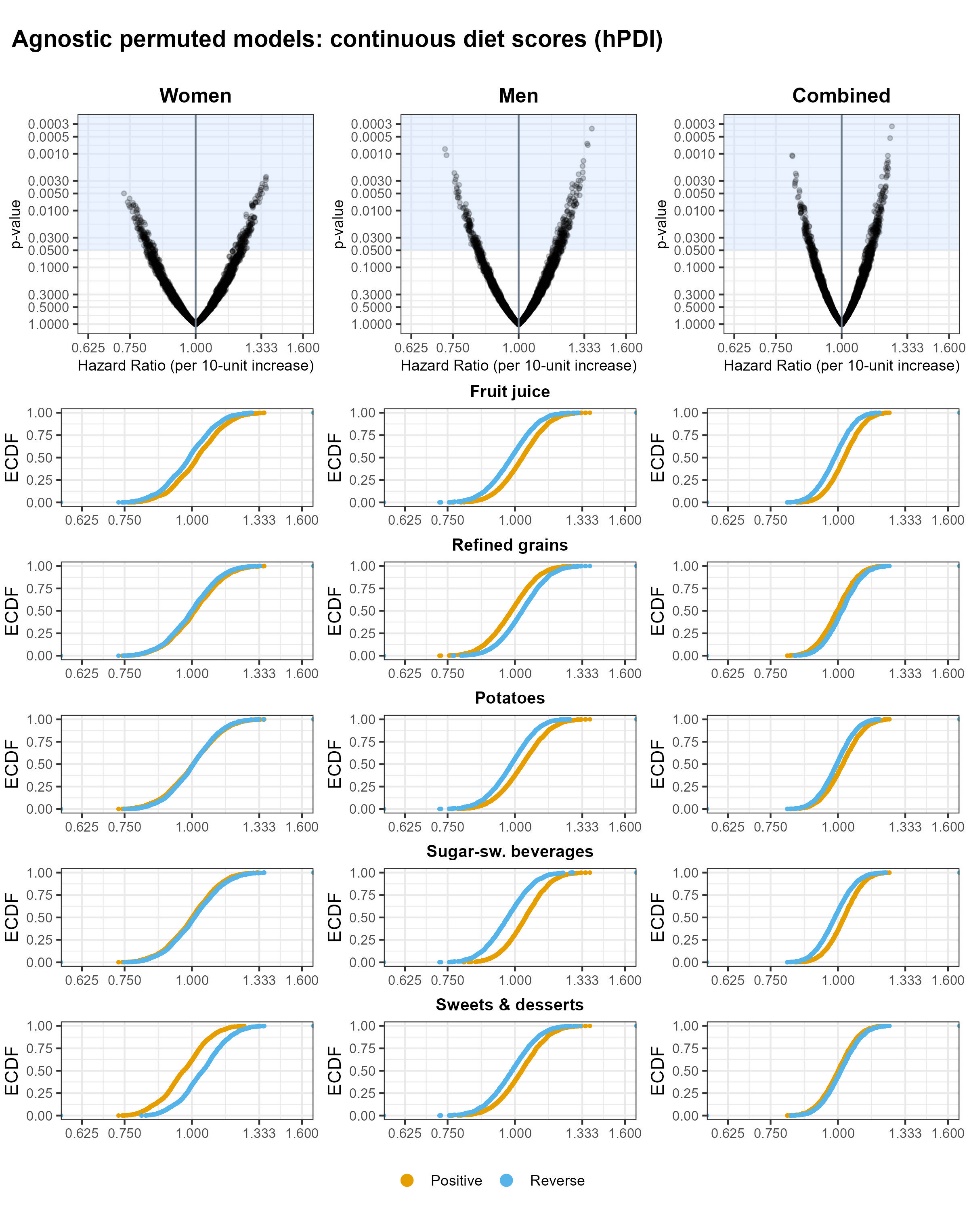

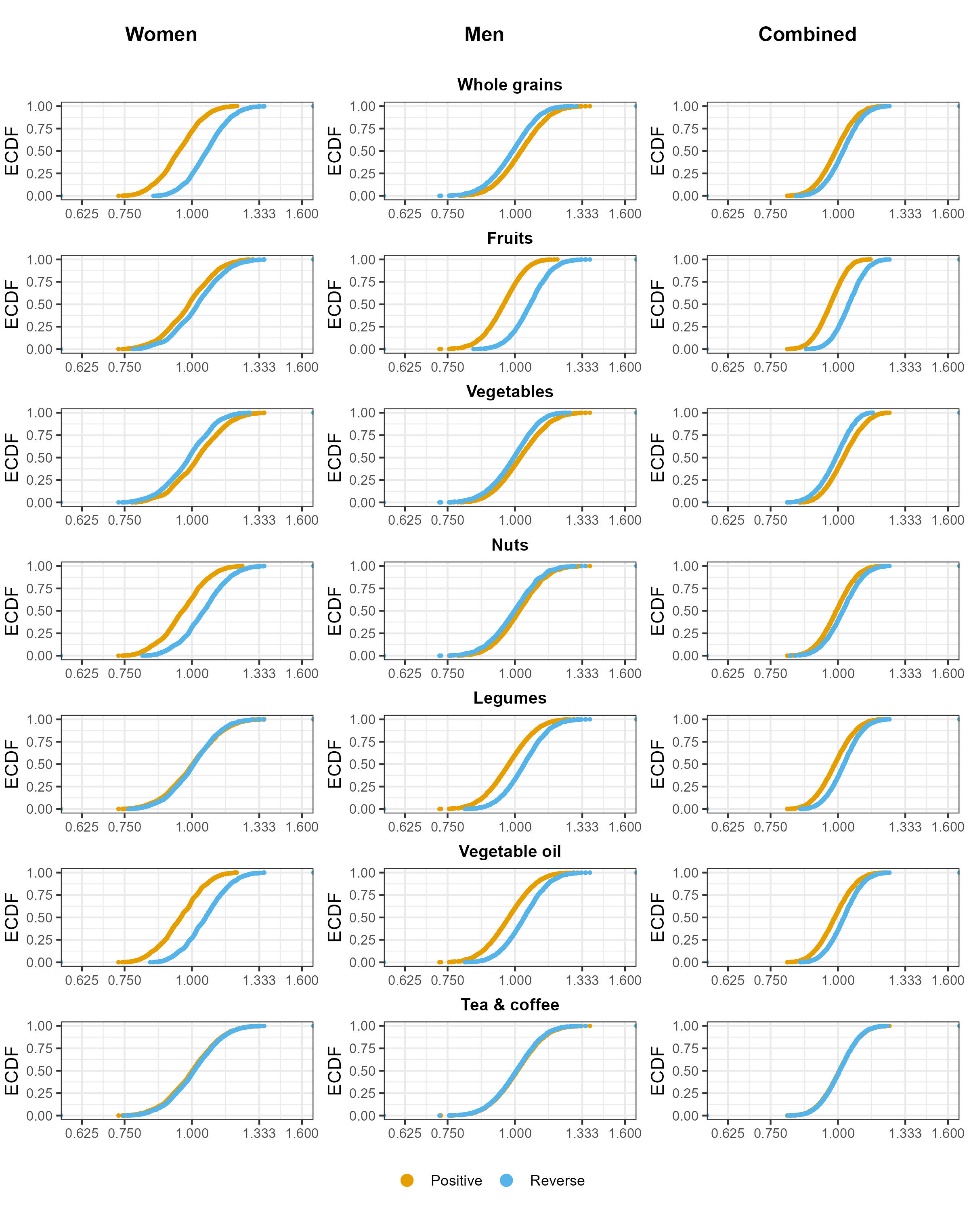

**Supplementary Figure 6.** Agnostic permutated models where each of the 12 plant-based food groups are either positively (i.e., more consumption is better; depicted in orange) or reversely (less consumption is better, depicted in blue) scored, using all combinations (empirical cumulative density function (ECDF) of hazard ratios for highest deciles compared to lowest decile of hPDI score).

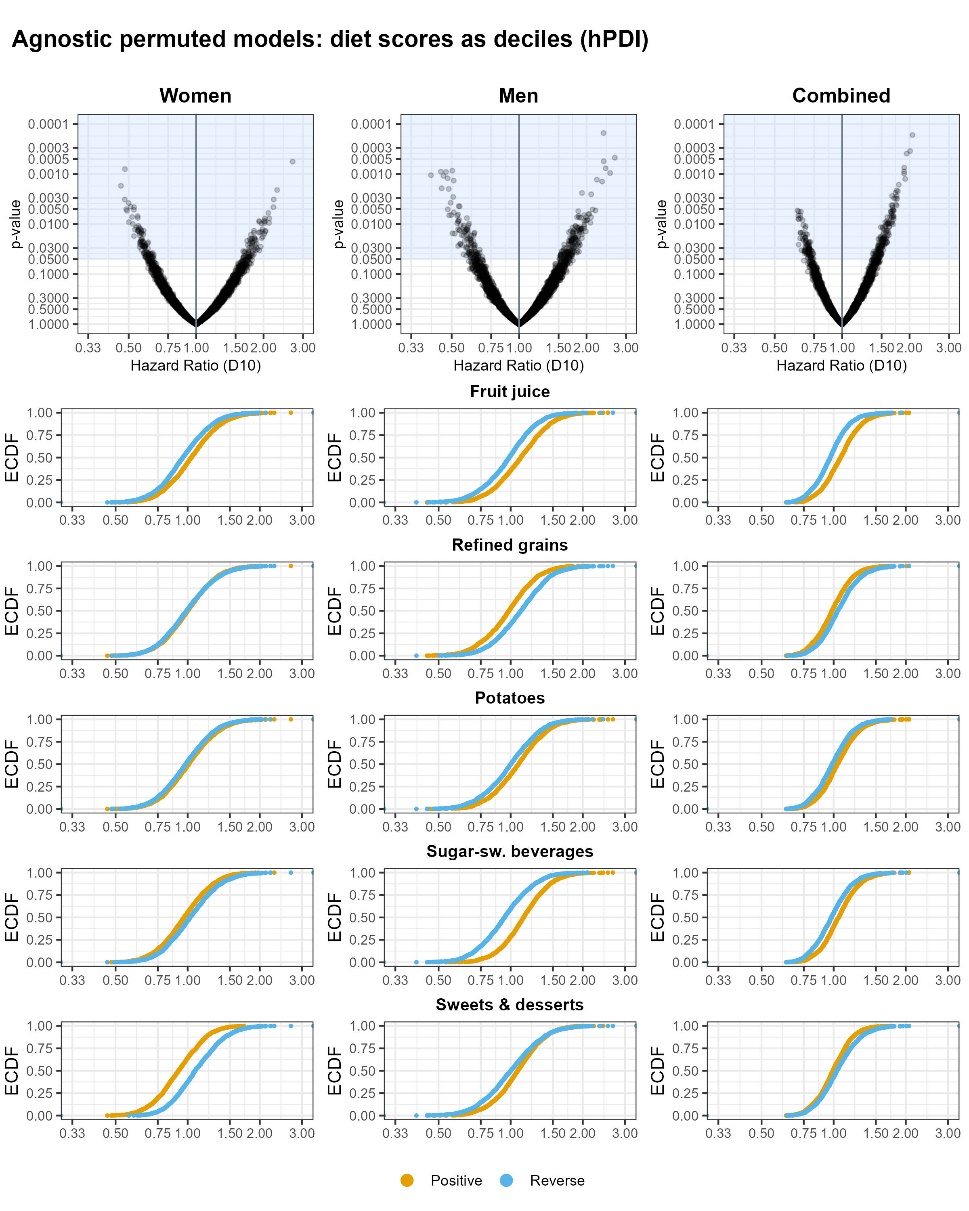

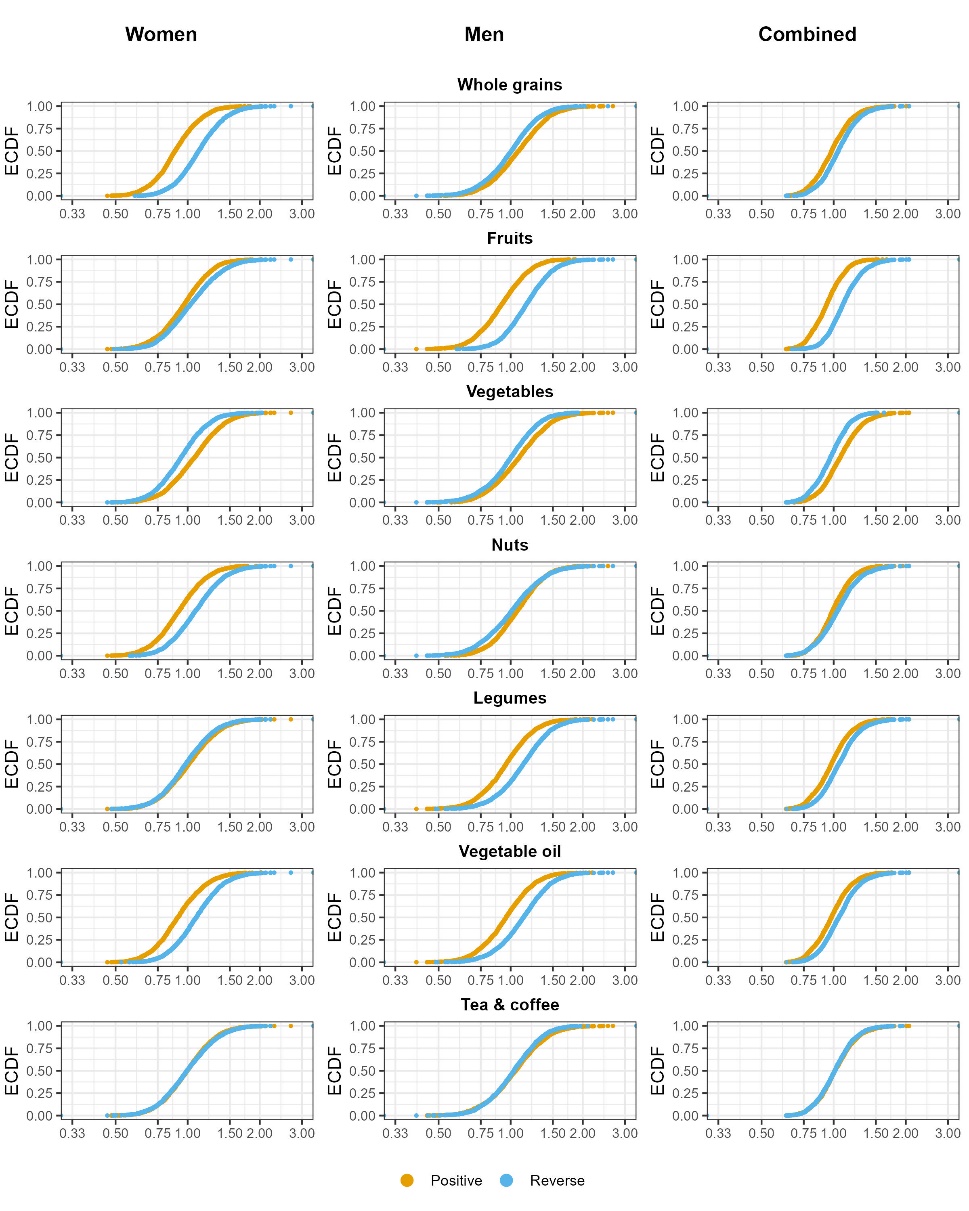
